# Landscape of blood group antigens and alleles in the Indian population from whole genome sequences

**DOI:** 10.1101/2023.09.26.23296145

**Authors:** Mercy Rophina, Rahul C Bhoyar, Mohamed Imran, Vigneshwar Senthivel, Mohit Kumar Divakar, Anushree Mishra, Bani Jolly, Sridhar Sivasubbu, Vinod Scaria

## Abstract

Blood group antigens are genetically inherited macromolecular structures which form the underlying factor for inter individual variations in human blood. Currently there exists over 390 human blood group antigens corresponding to 44 blood group systems and 2 erythroid specific transcription factors. Distribution of these blood group antigens have been found to differ significantly among various ethnic populations. To date, there is a lack of comprehensive research that offers extensive blood group profiles for the Indian population. Whole genome sequence data (hg38) of 1029 self-declared healthy Indian individuals generated as a part of the pilot phase IndiGen programme were used for the analysis. Variants spanning the genes of 44 blood group systems and two transcription factors KLF1, GATA1 were fetched and annotated for their functional consequences. Our study reports a total of 40712 blood group related variants of which 695 were identified as non-synonymous variants in the coding region. Of the total non-synonymous variants, 105 were found to have a known blood phenotype. A total of 24 variants belonging to 12 blood groups were predicted to be deleterious by more than three computational tools. Our study was also able to identify a few rare blood phenotypes including Au(a-b+), Js(a+b+), Di(a+b-), In(a+b-) and KANNO-. This study is the first to use genomic data to understand the blood group antigen profiles of the Indian population, and it also systematically compares these profiles with those of other global populations.

**Key points:**

- Accurate characterization of the genomic landscape of known and rare blood group alleles and antigens in the Indian population using the whole genome sequencing data of 1029 self-declared healthy individuals
- Understanding the distinct similarities and differences in blood group genotypes and phenotypes across diverse global populations through systematic comparison of genomic datasets.

**Graphical abstract:** 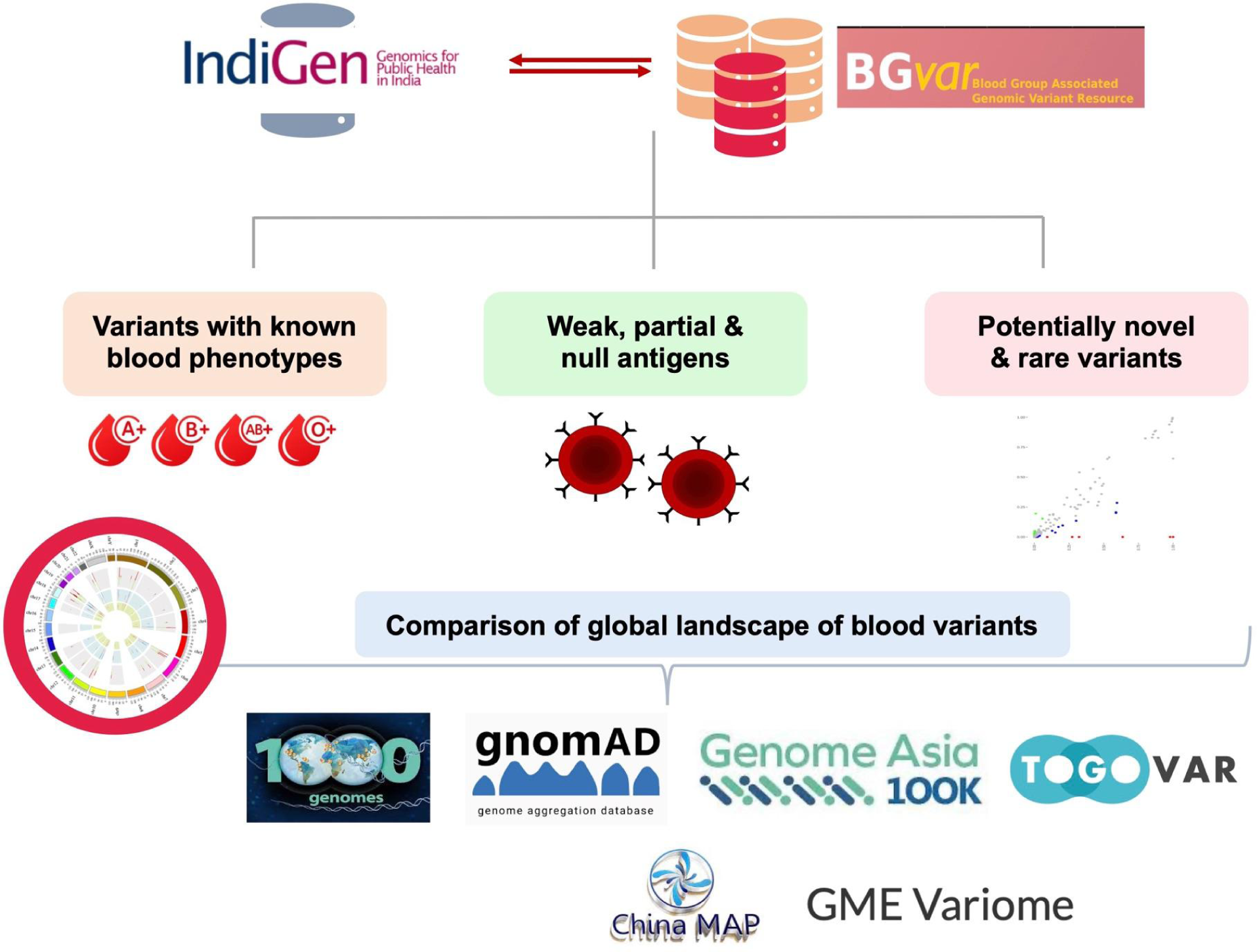

## Introduction

Blood group antigens are genetically inherited macromolecular structures which form the underlying factor for inter individual variations in human blood. These antigens are primarily protein, glycolipids or glycoproteins (Dean, 2005). With constant increase in demand for blood and blood related products, characterization of human blood groups has been a routine practice in transfusion science and in blood related disorders. Blood type generally represents the complete set of red cell/platelet surface antigens carried by an individual. Any mismatch or incompatibility between these antigens during transfusion procedures or pregnancies can lead to severe immune responses which can sometimes bring life threatening complications. In addition, human blood groups are found to have profound disease associations (Mitra et al., 2014)(Tufano et al., 2013),(Hiltunen et al., 2009),(D.-S. Wang et al., 2012),(Gates et al., 2011),(Anstee, 2010),(“Complications of Blood Transfusion,” 2006)

Currently there exists about 360 human blood group antigens corresponding to 41 blood group systems and 2 erythroid specific transcription factors(Storry et al., 2019). Distribution of these blood group antigens have been found to differ significantly among various ethnic populations. Knowing the indigenous prevalence of blood groups is vital for systematic functioning of blood bank units for effectively managing alloimmunization and multi-transfusion cases. Recent years have witnessed numerous studies that have extensively performed population scale blood phenotype frequency analysis (E.f.b.t.m. et al., 2018),(AlBilali. et al., 2017),(AlSuhaibani et al., 2015), (Balci, 2010), (Yu et al., 2016),(Owaidah et al., 2020),(Bashwari et al., 2001),(Sarhan et al., 2009),(Canizalez-Román et al., 2018), (Nazli et al., 2015),(Doku et al., 2019),(Golassa et al., 2017)

India being an abode of over 4500 diverse ethnic groups with the second largest population density of 1.3 billion, demands the need to maintain indigenous population specific blood group profiles for effective regulation of transfusion practices (Mastana, 2014). There exists a handful of regional population studies in India, revealing the prevalence of major blood groups - ABO and RH (Garg et al., 2014),(Singh et al., 2016),(Chandra & Gupta, 2012),(Kaur et al., 2013),(Kaur et al., 2013; Mondal et al., 2012),(Periyavan et al., 2010),(Nanu & Thapliyal, 1997).Although ABO and RH blood groups play predominant roles in immunohematological procedures, a range of minor human blood group systems with significant impact in transfusion have been exposed in the last decade. In developing countries like India alloimmunization complications due to clinically significant minor blood groups are not completely considered. Recipient groups including young women, pregnant females and patients expected to undergo multiple transfusion procedures because of blood related disorders require routine testing for significant minor blood group antigens. Very minimal studies exist, which have looked at the landscape of clinically significant minor blood group antigens in the nation. (Lamba et al., 2013), (Thakral et al., 2007).

Owing to the diversity of human blood group antigens, obtaining a complete blood group profile using classical serological or molecular techniques becomes impractical. An alternative approach of utilizing the whole genome sequence data of an individual to retrieve the complete blood profile was demonstrated by Lane et al in 2016 (Lane et al., 2016). Large scale genomic datasets have allowed researchers to explore *‘previously neglected antigens’*. One such discovery was reported by Moller et al in 2016, where a 5821 bp deletion encompassing exons 5,6, and 7 of the ABO gene was found in 20 individuals primarily of African origin in the 1000 Genomes project data (Möller et al., 2016).

Our study aims to systematically assess large scale genomic data to explore complete blood group antigen profiles of Indian indigenous population of the nation thereby paving way to understand the geographical and ethnic distribution of blood group antigens and associated phenotypes.

## Materials and methods

### Reference and study datasets

Reference dataset includes a comprehensive list of ISBT approved blood group related variants, documented in a pre-formatted template. Whole genome sequence data (hg38) of 1029 self-declared healthy Indian individuals generated as a part of the pilot phase IndiGen programme were used for the analysis. The study dataset comprised a total of 59646267 genetic variants which includes single-nucleotide variants (SNVs), insertions and deletions. Comprehensive list of genetic variations was retrieved in the variant call format (VCF) files and were used for further analyses. In addition, a range of large global population scale datasets including the 1000 genomes project, gnomAD, GenomeAsia 100K, Greater Middle Eastern Variome, Qatar, Iranome, Singapore Sequencing Malay Project (SSMP), Singapore Sequencing Indian Project (SSIP), China Metabolic Analytics Project (ChinaMAP) and Japanese genome variations (TogoVar) were used for comparison analysis.

### Processing and annotation of variants

There exist 53 unique genes associated with human blood groups. In the primary level of data processing, all the genetic variants spanning human blood group genes and erythroid specific transcription factors were filtered. **Supplementary Table 1.** provides the list of genes along with their corresponding locus genomic reference (LRG) coordinates. Filtered variants were subsequently annotated for a range of functional consequences including SIFT (Ng & Henikoff, 2003), Polyphen (Adzhubei et al., 2013) LRT, MutationTaster, Mutation Assessor (Gnad et al., 2013), FATHMM (Shihab et al., 2013), PROVEAN (Choi & Chan, 2015), CADD (Rentzsch et al., 2019), GERP (Davydov et al., 2010), PhyloP (Pollard et al., 2010) and PhastCons (Siepel et al., 2005) using Annotate Variation (ANNOVAR) (K. Wang et al., 2010)). A schematic representation of the methodology is depicted in **Figure 1**.

**Figure 1.**
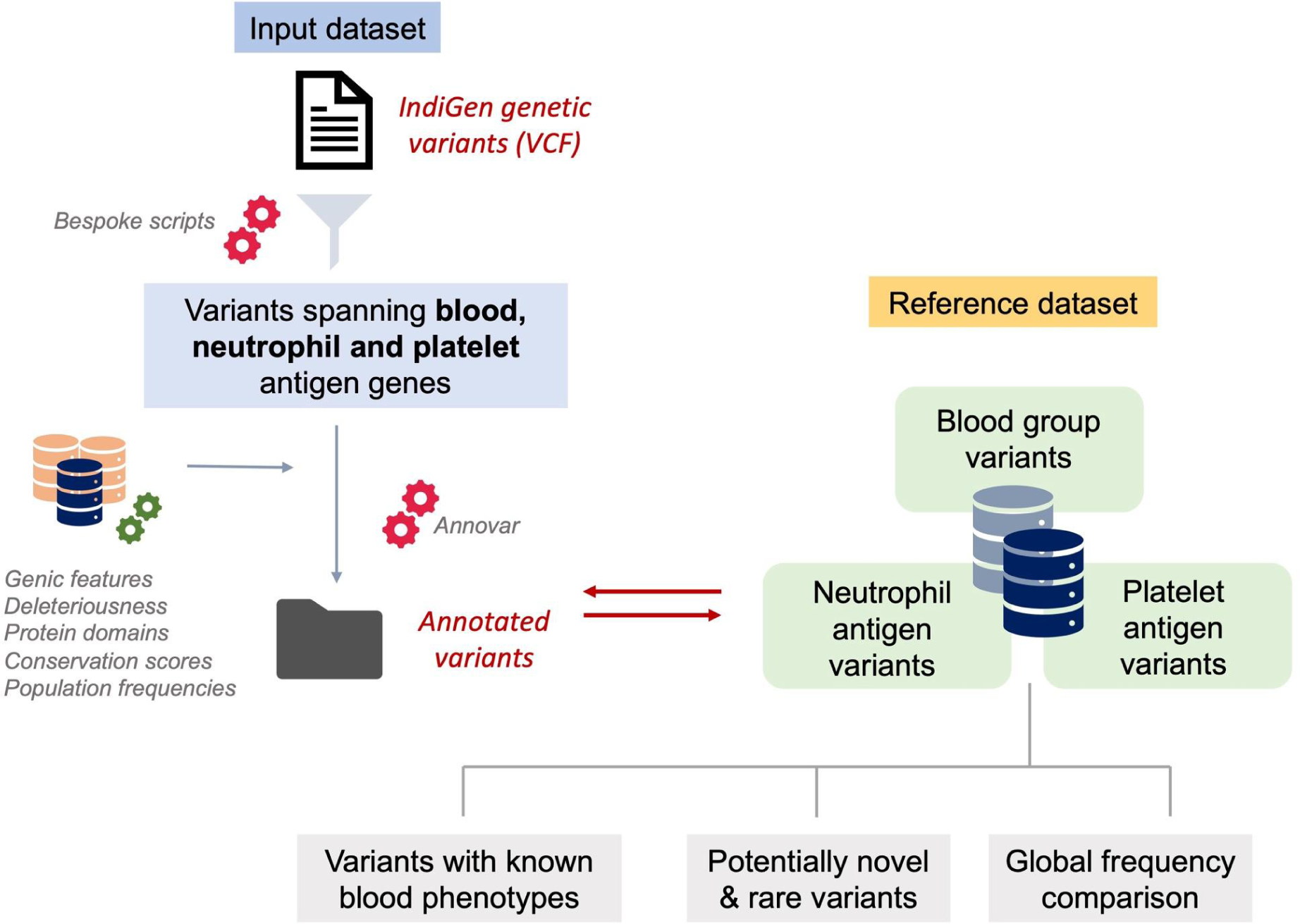
Schematic representation of methodology followed in fetching human blood groups associated genetic variations in the Indian population.

### Identification of variants with known blood phenotypes in the Indian population

The annotated variants were systematically classified into two categories: those that are associated with a specific blood phenotype and those that are not. All the blood group systems with no blood group phenotype associated variants were conferred with the reference nomenclature as previously described {ref}.

### Identification of potentially novel and rare variants

In subsequent levels of data analysis, the filtered variants were systematically classified as potentially novel/rare variants based on their SNP identification number (dbSNP ID) and minor allele frequencies (MAF). Any filtered variant lacking a dbSNP ID and literature evidence were deemed as potentially novel. Exonic novel variants were further assessed for their deleteriousness predicted by a range of computational tools. In addition, variants with MAF < 1% with absence of reported blood phenotypes were deemed as rare variants.

### Estimation and comparison of blood group related allele frequencies among various global populations

To assess the distribution of blood group, neutrophil, and platelet variants in the Indian population, a comparative analysis was conducted by examining the allele frequencies of these variants across major global populations. The comparisons included data from diverse resources like 1000 Genomes project, Genome Aggregation Database (gnomAD), Genome Asia, Greater Middle Eastern Variome, Qatar Genomes and Exomes, Singapore Sequencing

Malay Project (SSMP), Singapore Sequencing Indian Project (SSIP), China Metabolic Analytics Project (ChinaMAP) and Japanese genome variations (TogoVar)

### Statistical analysis

Significantly distinct blood group, neutrophil and platelet antigen alleles specific for the Indian population were identified using Fisher’s exact test with a P-value < 0.05. The filtered alleles/variants were subsequently checked for their clinical relevance in transfusion settings.

## Results

### Human blood group associated variants and annotations

A total of 40712 variants spanning 44 blood group genes were filtered from the preliminary level of analysis. Detailed tabulation of the variant counts in each gene is provided in **Supplementary Table 2.** Functional classification of the filtered variants revealed a total of 1197 variants spanning the exonic and splice site regions, out of which 695 were found to be non-synonymous 398 were synonymous, 1 were stop loss and 14 were stop gain variants as shown in **Supplementary Figure 1(A and B).**

### Overview of blood group variants in India

A total of 29, 17 and 4 variants mapping back to ABO, RHD and RHCE genes were found to possess known blood phenotypes. In addition, 68 variants were identified spanning 27 other blood genes corresponding to 25 systems. Comprehensive summary of the list of identified blood variants is listed in **Table 1**. Out of the 44 blood group systems and 2 erythroid specific transcription factors 22 systems had no variant associated with blood group phenotype and therefore they were conferred with the reference nomenclature. Identified blood group phenotypes for all the systems are tabulated in **Table 2**.

**Table 1.** Summary of blood group variants with known blood phenotypes identified from the study dataset.

| Chr | Position | Ref Allele | Alt Allele | Allele Count | Allele Frequency | Allele Number | Blood Group | Gene Name | Blood Phenotype |
| --- | --- | --- | --- | --- | --- | --- | --- | --- | --- |
| 7 | 30912043 | C | T | 9 | 0.004 | 2052 | COLTON | AQP1 | CO:2 or Co(b+) |
| 7 | 30912049 | A | G | 1 | 0 | 2052 | COLTON | AQP1 | CO:1;2;3;4 |
| 1 | 207322436 | G | C | 1 | 0 | 2048 | CROMER | CD55 | CROM:3 or Tc(b+) |
| 1 | 207326820 | C | T | 4 | 0.002 | 2050 | CROMER | CD55 | CROM:-12 or SERF- |
| 17 | 44251253 | G | A | 12 | 0.006 | 2050 | DIEGO | SLC4A1 | DI:1;-2 or Di(a+b-) |
| 12 | 14840624 | A | T | 3 | 0.001 | 2048 | DOMBROC<br>K | ART4 | DO:-8 or DOLG- |
| 1 | 159205564 | G | A | 665 | 0.324 | 2050 | DUFFY | ACKR1 | FY:2 or Fy(b+) |
| 1 | 159205704 | C | T | 12 | 0.006 | 2050 | DUFFY | ACKR1 | Fy(a+w) |
| 1 | 159205834 | G | A | 2 | 0.001 | 2052 | DUFFY | ACKR1 | Fy(a-b-) |
| 9 | 133154258 | G | T | 92 | 0.045 | 2052 | FORS | GBGT1 | FORS:-1 (FORS-) |
| 9 | 133162355 | G | A | 718 | 0.358 | 2006 | FORS | GBGT1 | FORS:-1 (FORS-) |
| 2 | 126693891 | C | T | 2 | 0.001 | 2052 | GERBICH | GYPC | GE:-10 or GEPL- |
| 3 | 161086379 | C | T | 263 | 0.129 | 2044 | GLOB | B3GALN<br>T1 | GLOB:1 (P+) |
| 19 | 48750557 | A | C | 7 | 0.003 | 2052 | H | FUT1 | H- |
| 19 | 48751247 | G | A | 286 | 0.139 | 2052 | H | FUT1 | H+ |
| 6 | 10586494 | G | A | 5 | 0.002 | 2048 | I | GCNT2 | I+W |
| 11 | 35176644 | G | C | 28 | 0.014 | 2050 | IN | CD44 | In(a+b-) |
| 11 | 35189886 | C | A | 1 | 0 | 2050 | IN | CD45 | IN:-4 or INJA- |
| 4 | 88092344 | C | T | 32 | 0.016 | 2046 | JUNIOR | ABCG2 | Jr(a+w) |
| 4 | 88118244 | G | A | 1 | 0 | 2052 | JUNIOR | ABCG2 | Jr(a-) |
| 4 | 88131171 | G | T | 236 | 0.115 | 2046 | JUNIOR | ABCG2 | Jr(a+w) |
| 20 | 4699875 | G | A | 97 | 0.047 | 2054 | KANNO | PRNP | KANNO- |
| 7 | 142943026 | A | G | 1 | 0 | 2052 | KELL | KEL | KEL:6,-7 or Js(a+b-) |
| 7 | 142946304 | C | T | 1 | 0 | 2048 | KELL | KEL | KEL:-26 or TOU- |
| 7 | 142953916 | G | A | 3 | 0.001 | 2050 | KELL | KEL | KEL:-22 |
| 7 | 142954267 | G | A | 2 | 0.001 | 2052 | KELL | KEL | KEL:3,-4,-21 or Kp(a+b-c-) |
| 7 | 142954320 | C | T | 1 | 0 | 2050 | KELL | KEL | Kmod |
| 7 | 142957921 | G | A | 14 | 0.007 | 2052 | KELL | KEL | KEL:1,-2 or K+k- |
| 7 | 142960939 | C | T | 4 | 0.002 | 2046 | KELL | KEL | KEL:-18 |
| 7 | 142960940 | G | A | 5 | 0.002 | 2046 | KELL | KEL | KEL:-18 |
| 18 | 45730450 | G | A | 625 | 0.305 | 2052 | KIDD | SLC14A1 | Jk(a+W) |
| 18 | 45731089 | G | A | 1 | 0 | 2048 | KIDD | SLC14A1 | Jk(a+W) |
| 18 | 45748385 | C | T | 18 | 0.009 | 2050 | KIDD | SLC14A1 | JK:-3 or Jk(a-b-) |
| 19 | 12885926 | A | G | 1016 | 0.498 | 2040 | KLF1 | KLF1 | Obsolete Normal BG<br>phenotype |
| 1 | 207587428 | C | T | 597 | 0.292 | 2048 | KNOPS | CR1 | KN:5 or Yk(a-) |
| 1 | 207609424 | G | A | 10 | 0.005 | 2050 | KNOPS | CR1 | KN:2 or Kn(b+) |
| 2 | 219213284 | C | T | 1 | 0 | 2050 | LANGEREIS | ABCB6 | Lan weak |
| 2 | 219216123 | C | T | 8 | 0.004 | 2054 | LANGEREIS | ABCB6 | Lan weak |
| 2 | 219216694 | G | A | 5 | 0.002 | 2048 | LANGEREIS | ABCB6 | Lan weak |
| 2 | 219217783 | G | A | 2 | 0.001 | 2046 | LANGEREIS | ABCB6 | Lan- |
| 19 | 44812188 | G | A | 2 | 0.001 | 2048 | LUTHERAN | BCAM | LU:1 or Lu(a+) |
| 19 | 44813447 | T | A | 2 | 0.001 | 2048 | LUTHERAN | BCAM | LU:-8,14 |
| 19 | 44814191 | C | T | 7 | 0.003 | 2052 | LUTHERAN | BCAM | LU:-6,9 |
| 19 | 44819367 | C | T | 5 | 0.002 | 2046 | LUTHERAN | BCAM | LU:-26, LUBI- |
| 19 | 44819487 | A | G | 442 | 0.216 | 2044 | LUTHERAN | BCAM | LU:-18,19 or Au(a-b+) |
| 22 | 42693843 | T | C | 674 | 0.331 | 2038 | P1PK | A4GALT | P1+/-; Pk+ |
| 6 | 49615692 | C | T | 1 | 0 | 2048 | RHAG | RHAG | Rhmod |
| 1 | 42830785 | G | A | 1 | 0 | 2054 | SCIANNA | ERMAP | SC:-7 or SCAN- |
| 1 | 42830851 | G | A | 1 | 0 | 2050 | SCIANNA | ERMAP | SC:2 or Sc2+ |
| 1 | 42830860 | C | G | 1 | 0 | 2050 | SCIANNA | ERMAP | SC:4 or Rd+ |
| 7 | 100893176 | G | T | 96 | 0.047 | 2050 | YT | ACHE<br>(YT) | YT:2 or Yt(b+) |
| 7 | 100893967 | C | T | 32 | 0.016 | 2048 | YT | ACHE<br>(YT) | YTEG- |
| 7 | 100894064 | C | T | 12 | 0.006 | 2044 | YT | ACHE<br>(YT) | YT:-4 or YTLI- |
| 7 | 100894132 | C | T | 1 | 0 | 2038 | YT | ACHE<br>(YT) | YT:-5 or YTOT |
| 1 | 207609586 | A | G | 966 | 0.47 | 2054 |  |  |  |
| 9 | 137031410 | C | T | 2 | 0.001 | 2040 |  |  |  |
| 19 | 5831566 | G | C | 43 | 0.021 | 2044 | LEWIS |  |  |
| 19 | 5843958 | G | A | 5 | 0.002 | 2028 | LEWIS |  |  |
| 19 | 5844108 | G | A | 6 | 0.003 | 2026 | LEWIS |  |  |
| 19 | 5844228 | T | C | 14 | 0.007 | 2040 | LEWIS |  |  |
| 1 | 25390874 | C | G | 211 | 0.103 | 2046 | RH | RHCE | RH:27 or cE RHCE*03 or<br>RHCE*cE |
| 1 | 25392026 | C | G | 1 | 0 | 2048 | RH | RHCE | RH:3 (E+ partial, weak to<br>neg) or RH:4 (c+ weak)<br>RHCE*03.04 |
|  |  |  |  |  |  |  |  |  | RHCE*cE.04<br>RHCE*cEIV |
| 1 | 25402721 | T | A | 4 | 0.002 | 2046 | RH | RHCE | Comb Mut |
| 1 | 25420665 | T | C | 10 | 0.005 | 2036 | RH | RHCE | RH:2 (C+ partial)<br>RH:5 (e+ partial)<br>RH:8 (C W +)<br>RH:-51(MAR-)<br>RHCE*O2.08.01<br>RHCE*Ce.08.01<br>RHCE*CeCW |
| 1 | 25272575 | C | T | 1 | 0.001 | 1916 | RH | RHD | Type 61 RHD*01W.61<br>RHD*weak D type61 |
| 1 | 25272595 | G | C | 4 | 0.002 | 1910 | RH | RHD | Normal D antigen<br>RHD*01.01 |
| 1 | 25284544 | G | C | 1680 | 0.867 | 1938 | RH | RHD | Del RHD*01EL.32<br>RHD*DEL32 |
| 1 | 25284589 | C | T | 1 | 0.001 | 1934 | RH | RHD | Comb Mut |
| 1 | 25284599 | G | A | 1 | 0.001 | 1934 | RH | RHD | Type 151 RHD* 01W.151<br>RHD*weak D type<br>151 |
| 1 | 25284611 | G | T | 2 | 0.001 | 1934 | RH | RHD | Type 152 RHD* 01W.152<br>RHD*weak D type<br>152 |
| 1 | 25284632 | C | T | 1 | 0.001 | 1930 | RH | RHD | Type 122 RHD* 01W.122<br>RHD*weak D type<br>122 |
| 1 | 25284633 | G | A | 1 | 0.001 | 1930 | RH | RHD | Type 85 RHD* 01W.85<br>RHD*weak D type<br>85 |
| 1 | 25300956 | A | C | 2 | 0.001 | 1906 | RH | RHD | DFW RHD*18<br>RHD*DFW |
| 1 | 25300979 | G | A | 5 | 0.003 | 1908 | RH | RHD | Type 33 RHD*01W.33<br>RHD*weak D type<br>33 |
| 1 | 25301533 | G | C | 16 | 0.008 | 1904 | RH | RHD | Type 154 RHD* 01W.154<br>RHD*weak D type<br>154 |
| 1 | 25301552 | T | G | 11 | 0.006 | 1906 | RH | RHD | DFV RHD*08.01 |
|  |  |  |  |  |  |  |  |  | RHD*DFV |
| 1 | 25301597 | G | A | 2 | 0.001 | 1908 | RH | RHD | Comb Mut |
| 1 | 25303436 | G | A | 1 | 0.001 | 1910 | RH | RHD | Type 66 RHD*01W.66<br>RHD*weak D type 66 |
| 1 | 25303439 | G | A | 1 | 0.001 | 1910 | RH | RHD | Type 8 RHD*01W.8<br>RHD*weak D type 8 6 |
| 1 | 25303452 | A | G | 1 | 0.001 | 1908 | RH | RHD | Comb Mut |
| 1 | 25306881 | C | A | 31 | 0.016 | 1912 | RH | RHD | Comb Mut |
| 9 | 133255669 | CG | C | 64 | 0.031 | 2040 | ABO | ABO | B weak ABO*BW.18 |
| 9 | 133255801 | C | T | 507 | 0.248 | 2048 | ABO | ABO | Comb Mut |
| 9 | 133255902 | C | T | 462 | 0.226 | 2048 | ABO | ABO | Comb Mut |
| 9 | 133255928 | C | G | 503 | 0.245 | 2050 | ABO | ABO | Comb Mut |
| 9 | 133255929 | C | T | 11 | 0.005 | 2050 | ABO | ABO | Comb Mut |
| 9 | 133255935 | G | T | 502 | 0.245 | 2050 | ABO | ABO | Comb Mut |
| 9 | 133255960 | G | A | 458 | 0.224 | 2048 | ABO | ABO | Comb Mut |
| 9 | 133255963 | G | T | 35 | 0.017 | 2048 | ABO | ABO | Comb Mut |
| 9 | 133256028 | C | T | 506 | 0.247 | 2052 | ABO | ABO | Comb Mut |
| 9 | 133256042 | C | T | 1 | 0 | 2052 | ABO | ABO | A x /A weak<br>ABO*AW.30.01 or B 3<br>ABO*B3.02 |
| 9 | 133256050 | C | T | 463 | 0.226 | 2052 | ABO | ABO | Comb Mut |
| 9 | 133256074 | G | A | 504 | 0.246 | 2050 | ABO | ABO | Comb Mut |
| 9 | 133256082 | G | A | 1 | 0 | 2050 | ABO | ABO | Comb Mut |
| 9 | 133256085 | A | T | 462 | 0.225 | 2050 | ABO | ABO | O ABO*0.06 O53 (O 6 ) |
| 9 | 133256091 | T | C | 1 | 0 | 2048 | ABO | ABO | Comb Mut |
| 9 | 133256136 | G | A | 3 | 0.001 | 2048 | ABO | ABO | Comb Mut |
| 9 | 133256152 | A | G | 4 | 0.002 | 2048 | ABO | ABO | Comb Mut |
| 9 | 133256189 | C | T | 51 | 0.025 | 2048 | ABO | ABO | A 1 ABO*A1.02 |
| 9 | 133256204 | C | T | 8 | 0.004 | 2048 | ABO | ABO | Comb Mut |
| 9 | 133256205 | G | C | 514 | 0.251 | 2048 | ABO | ABO | Comb Mut |
| 9 | 133256234 | GT | G | 19 | 0.009 | 2048 | ABO | ABO | Comb Mut |
| 9 | 133256264 | G | A | 108 | 0.053 | 2046 | ABO | ABO | Comb Mut |
| 9 | 133257486 | T | C | 973 | 0.475 | 2048 | ABO | ABO | Comb Mut |
| 9 | 133258116 | G | A | 467 | 0.228 | 2044 | ABO | ABO | Comb Mut |
| 9 | 133259833 | G | A | 457 | 0.222 | 2054 | ABO | ABO | B weak ABO*BW.26 |
| 9 | 133259834 | C | T | 458 | 0.223 | 2054 | ABO | ABO | O ABO*O.01.01 O01 (O 1) |
| 9 | 133261367 | C | A | 455 | 0.222 | 2046 | ABO | ABO | Comb Mut |
| 9 | 133261370 | C | T | 4 | 0.002 | 2046 | ABO | ABO | Comb Mut |
| 9 | 133262144 | C | A | 12 | 0.006 | 2050 | ABO | ABO | Comb Mut |
| 9 | 133257521 | T | TC | 886 | 0.4322 | 2050 | ABO | ABO | Comb Mut |
| 4 | 144120554 | C | A | 1187 | 0.5882 | 2018 | MNS | GYPA | Comb Mut |
| 4 | 144120555 | T | C | 1187 | 0.5882 | 2018 | MNS | GYPA | Comb Mut |
| 4 | 144120567 | A | G | 1206 | 0.5953 | 2026 | MNS | GYPA | Comb Mut |
| 4 | 143997559 | C | G | 610 | 0.2984 | 2044 | MNS | GYPB | MNS:4,5 (s+, U+) altered U/GPB |
| 4 | 143997580 | G | A | 3 | 0.0015 | 2046 | MNS | GYPB | Comb Mut |
| 4 | 143999413 | G | C | 1 | 0.0005 | 2044 | MNS | GYPB | Comb Mut |
| 4 | 143999443 | G | A | 634 | 0.3096 | 2048 | MNS | GYPB | MNS:3 or S+ |
| 4 | 144001256 | G | C | 1 | 0.0005 | 2046 | MNS | GYPB | MNS:21or Mv+ |

**Table 2.**
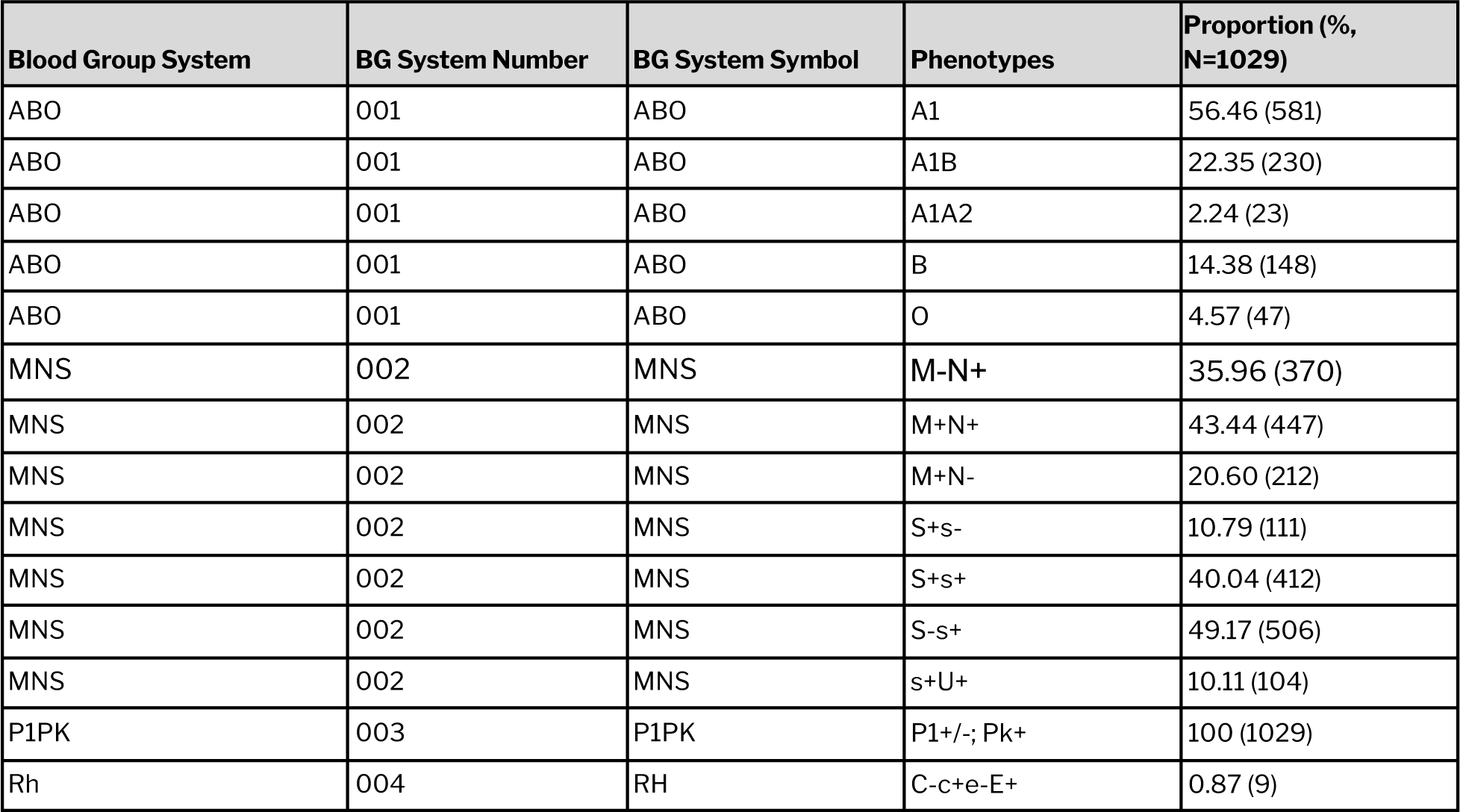
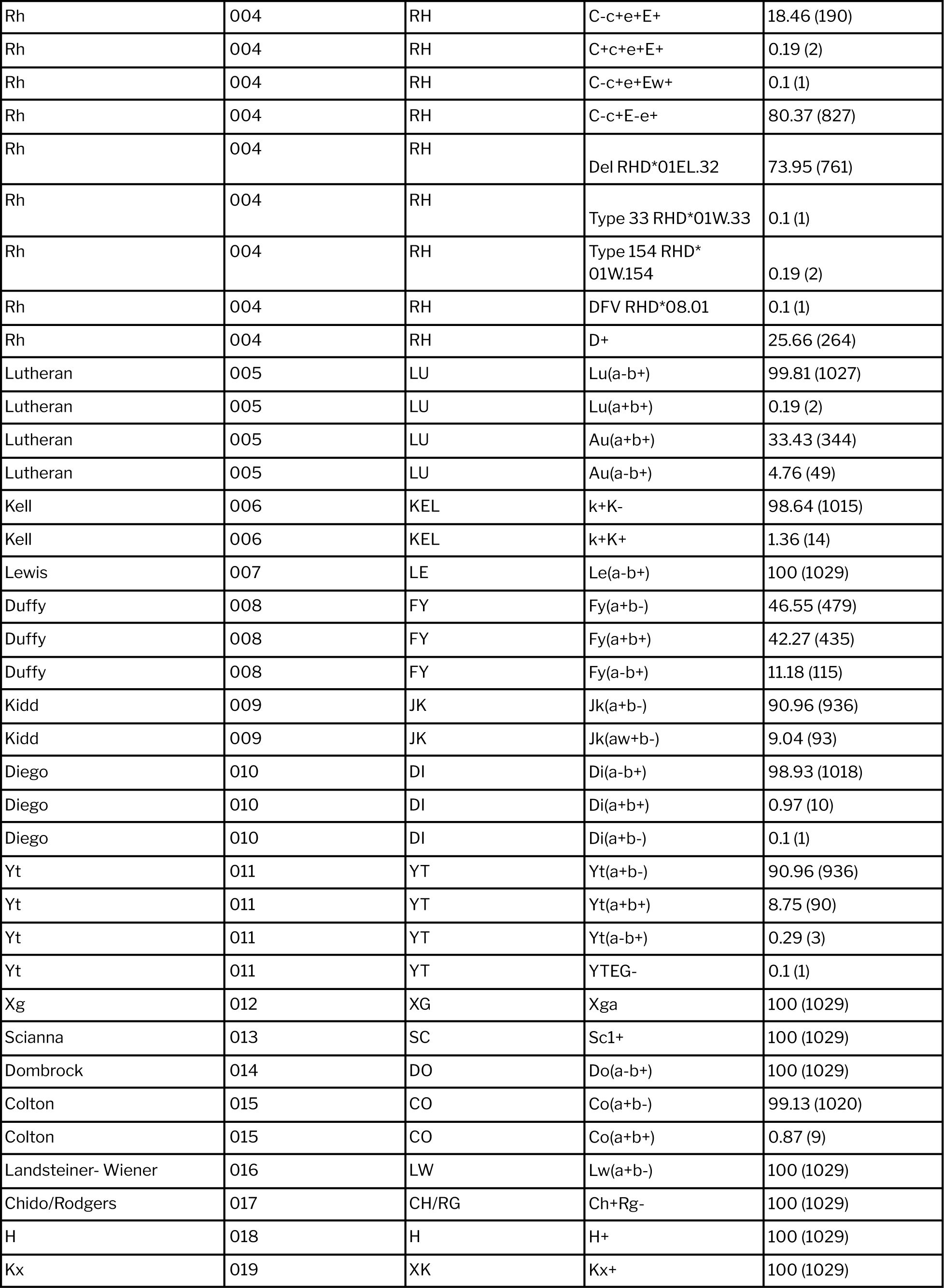
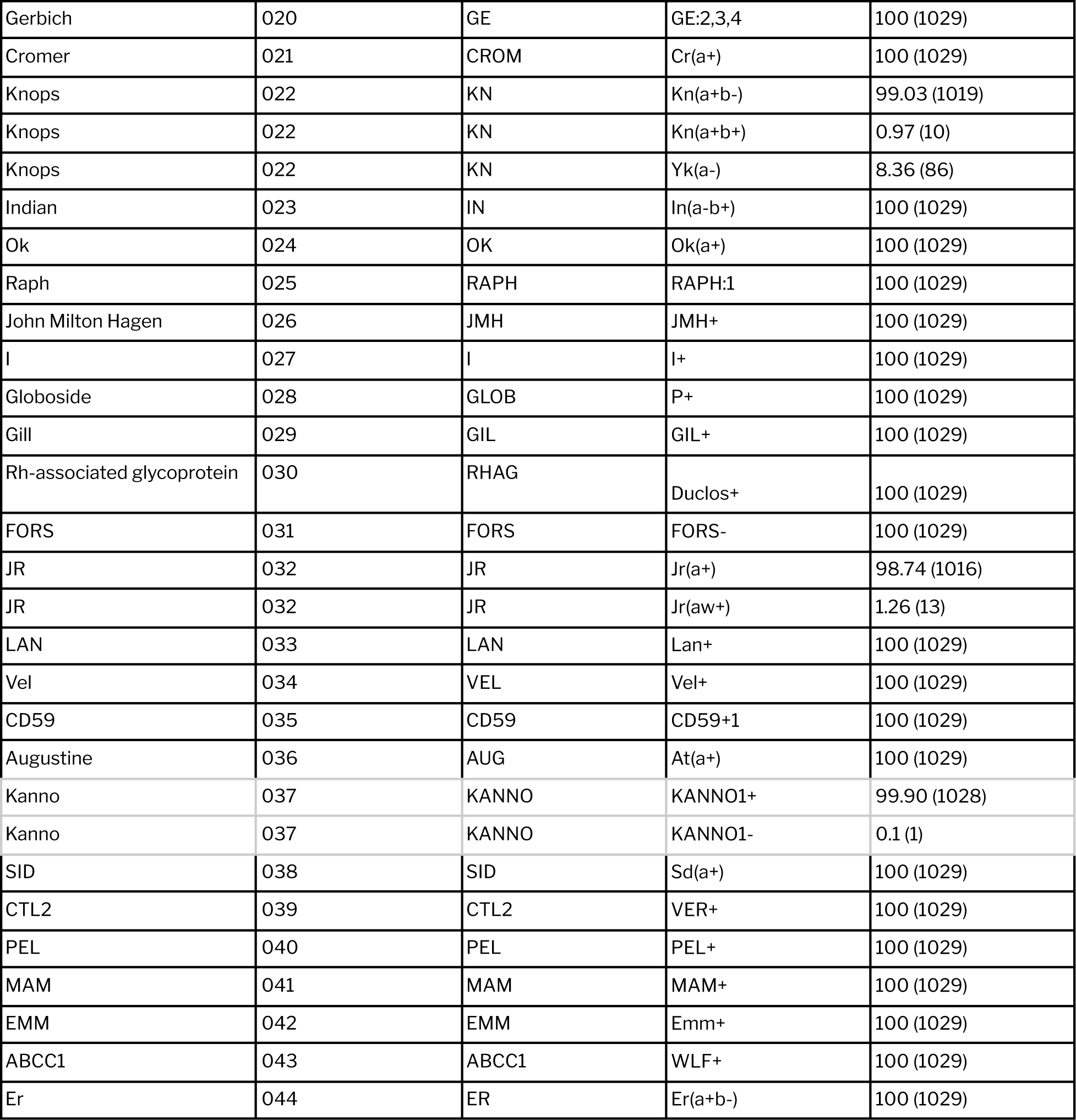
Tabulation of the blood phenotypes of all the 44 blood group systems identified in the Indian population.

### Overview of blood group phenotype

RH blood group is one of the highly polymorphic and severely immunogenic systems known till date. Although D antigen can be easily detected in most cases, variability of expression leads to weak expression of D antigens which are weakly immunogenic and are often missed during routine serological screening. Interestingly, we identified a rare partial D antigen, RHD*DFV (RHD*08.01, RHD*667G) which is well known to be associated with anti-D formation in carriers (Chen & Flegel, 2005; Fichou et al., 2018; Flegel et al., 2009; Grootkerk-Tax et al., 2006; Kappler-Gratias et al., 2014; Noizat-Pirenne et al., 2002). This RH phenotype carries a single nucleotide variation 667T>G which leads to amino acid change at position 223 (p.Phe223Val). It is reported that the amino acid site 223, which is located on the seventh transmembrane domain of the RHD protein plays an integral role in the formation of RHD epitopes. Evidence shows that change of amino acid at site 223 leads to the loss of 2 RHD epitopes (1.3 and 8.1) which subsequently contributes to the formation of alloanti-D antibodies if transfused with D+ RBCs (Grootkerk-Tax et al., 2006). Another RH weak antigen, RHD*DFV (RHD*18, RHD*497C) which results in moderate reduction of D antigen densities was also identified. Carriers of this antigen were found to have no anti-D antibody formation {ref}.

### Analysis of MNS blood group phenotype

The MNS blood group system is the second most complex after the RH system. MNS system consists of 49 antigens of which M/N, S/s/U are highly prevalent (Reid, 2009). It is well known that allo-anti U is one of the clinically significant antibodies associated with moderate to severe hemolytic reactions and HDFNs. {ref} A recent report from Francis LC et al., states that GYPB:c.251G>C change (p.Ser84Thr) is associated with an U-like allo antibody. This mutation which causes a change in amino acid site 84, located on the transmembrane region is found to alter the U/GPB protein (“27th Regional Congress of the ISBT, Copenhagen, Denmark, June 17-21, 2017,” 2017; Peyrard et al., 2012). MNS:4,5 (s+, U+) altered U/GPB phenotype is found in 10.11% of the Indian population. However, the clinical significance of this U-like alloantibody is still not clear.

### Potentially novel and rare blood group antigens

On comparing the allele frequencies of 695 exonic, non-synonymous blood variants identified in our study with global population datasets (1000 Genomes data) we found that 197 variants were found uniquely in the Indian population. About 315 and 134 variants were found at >2x and <2x frequencies respectively when compared to overall 1000 genomes population frequencies. And, 49 variants were found to be in a similar frequency range. Further, frequency distributions were also compared with 5 major subpopulations in 1000 genomes dataset namely Africans, Americans, Europeans, East Asians and South Asians. Although, as expected, variant frequencies in the Indian population dataset shared a high level of similarity with the South Asians, we were also able to fetch variants exclusive to our study dataset. A scatter plot depicting the distribution of variant frequencies across various global populations is depicted in **Figure 2**.

**Figure 2.**
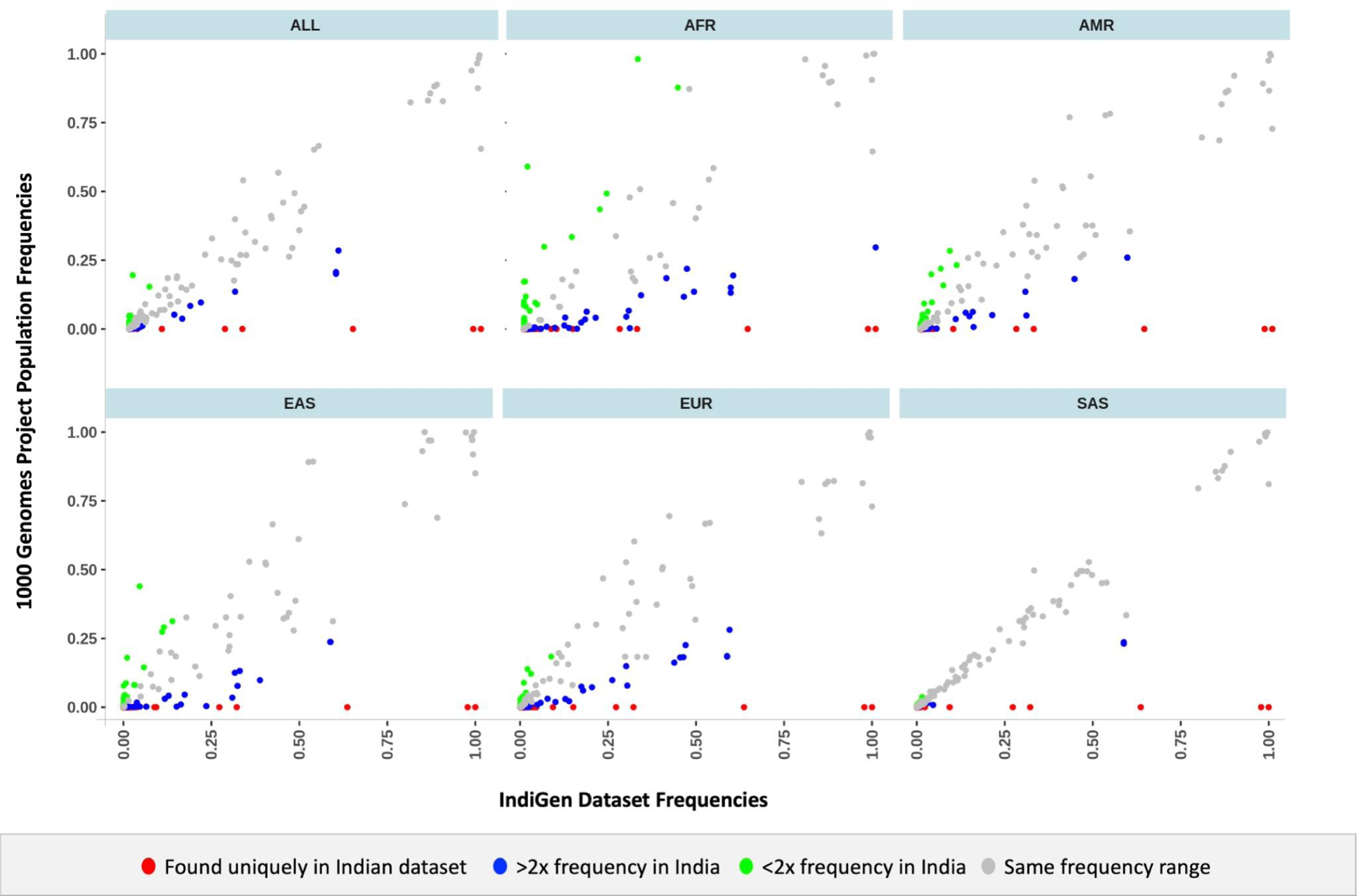
Scatter plot depicting the similarities and differences in the distribution of variant frequencies between Indian and various global populations.

Of the total variants, 143 exonic, non-synonymous variants which lacked dbSNP ID and literature evidence. These variants were found to span 37 unique blood genes and lacked any reported global population frequencies till date. Of these total variants, 24 variants belonging to 12 blood groups (LANGEREIS, GLOB, JUNIOR, CH/RG, COLTON, KELL, IN, DOMBROCK, PEL, DIEGO, BOMBAY and LEWIS) were predicted to be deleterious by more than three computational tools. Overview of the distribution of novel variants across various blood group genes is shown in **Supplementary Figure 1(C).** Interestingly, we observed a missense variant in FUT1 gene (p.Arg221Gly) which was predicted to have deleterious effects. An alternate variant reported at the same position (p.Arg221Cys) is well known to be associated with weak H phenotype (FUT1*01W.11)which thereby makes the identified FUT1:p.Arg221Gly a potentially novel weak H allele.

In addition, we also filtered a total of 588 exonic, non-synonymous variants with MAF<1% mapping back to 58 unique blood genes.

### Identification of weak/partial and null blood group antigens

A total of 25 variants associated with weak/partial and null blood phenotypes were identified in the study as listed in **Table 3**.

**Table 3.**
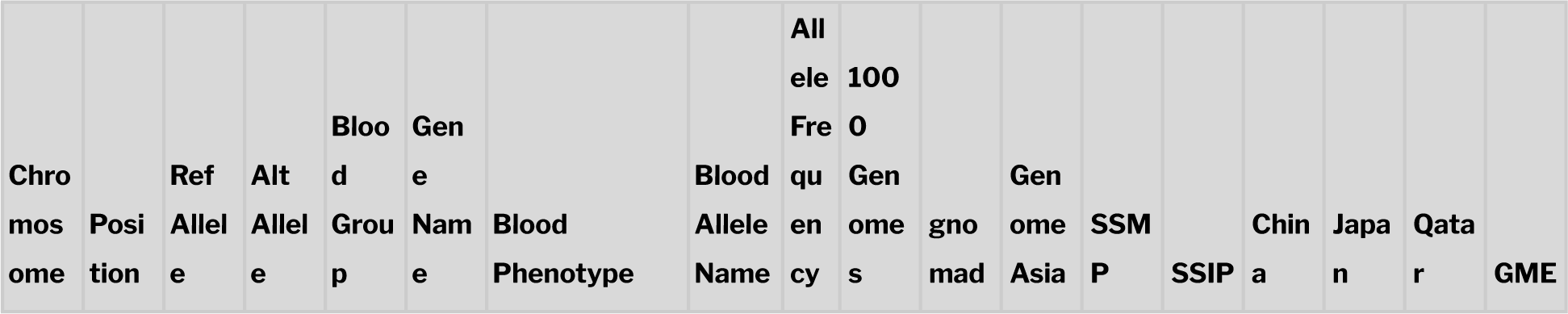
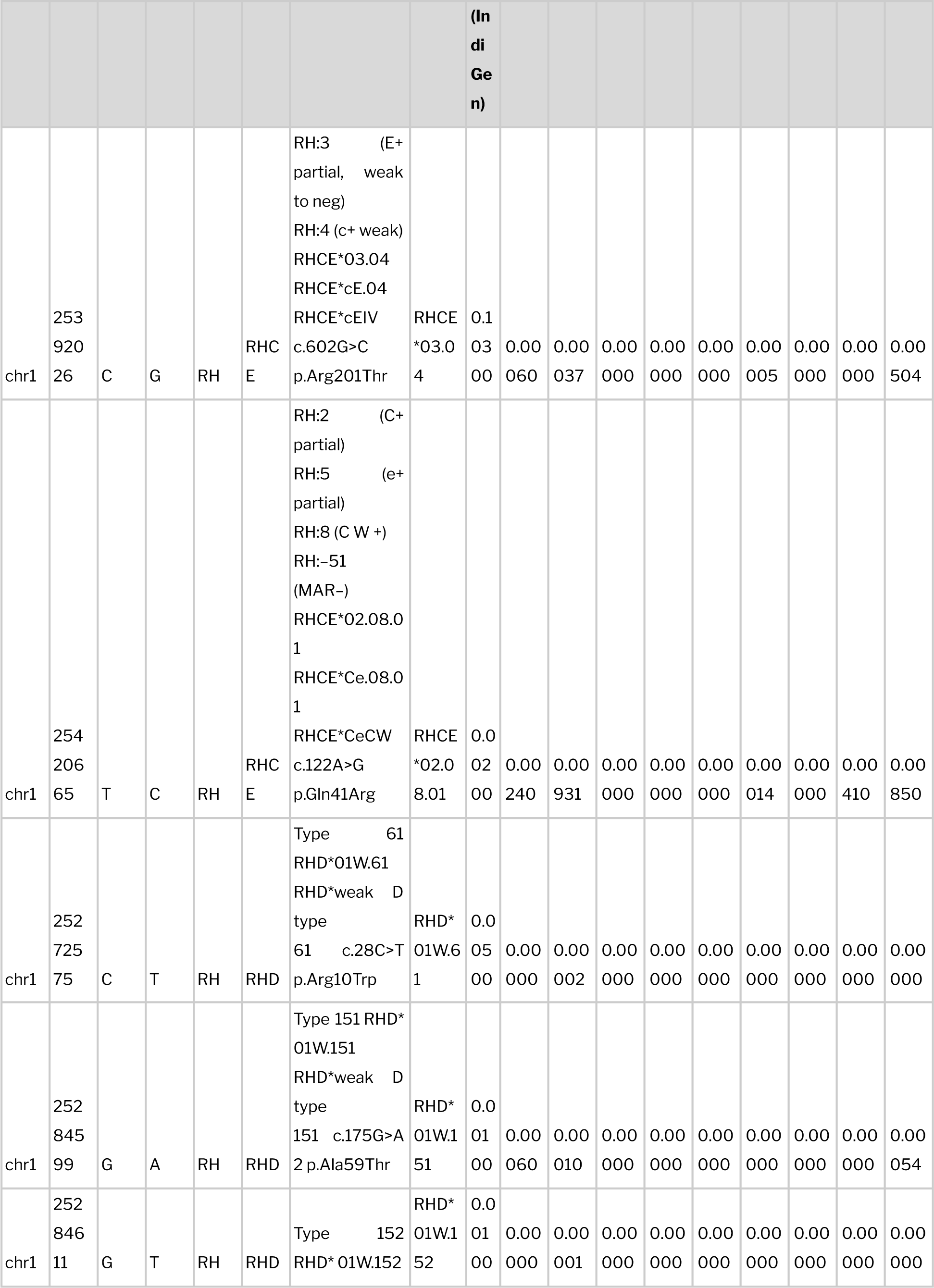
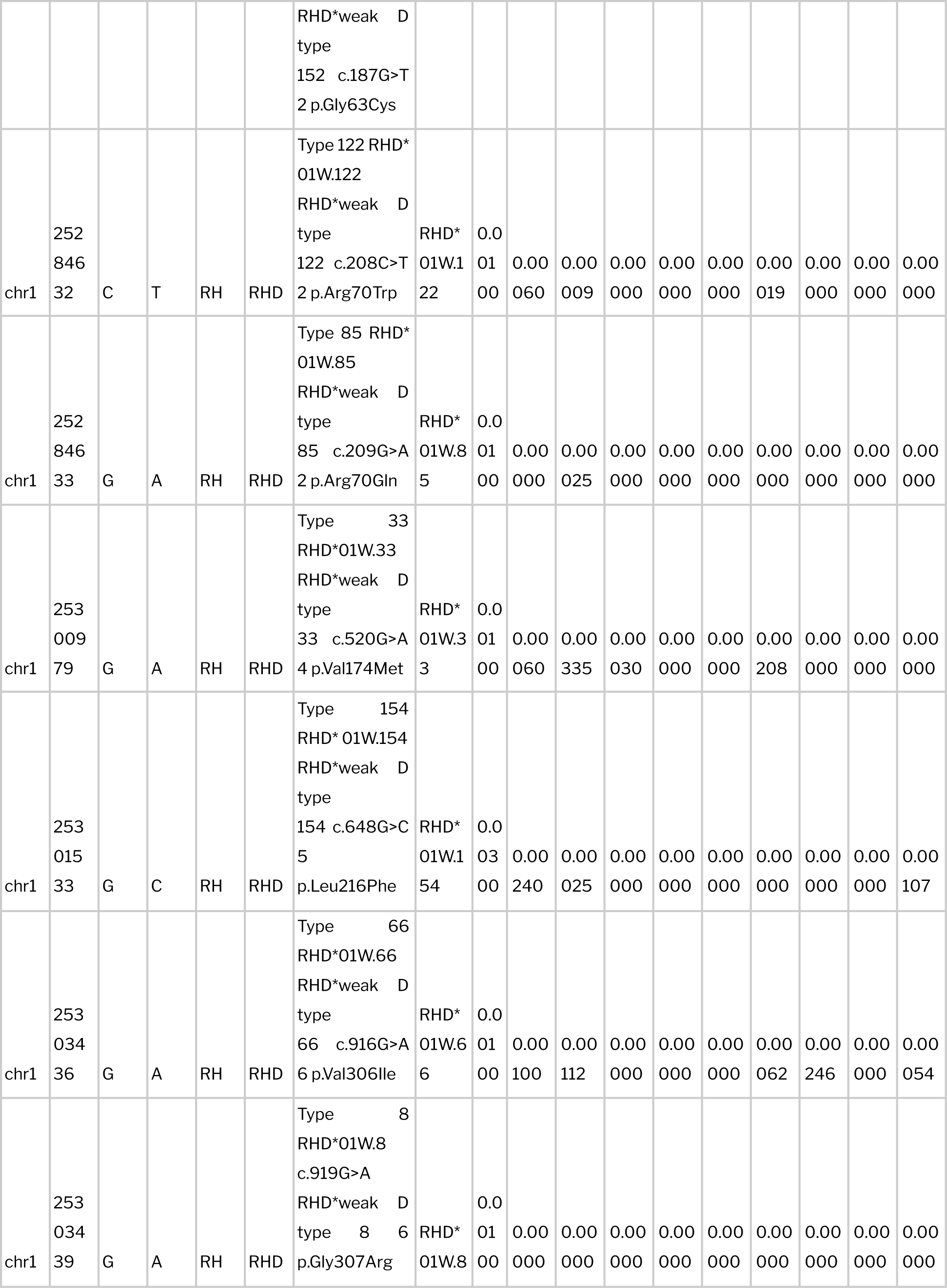
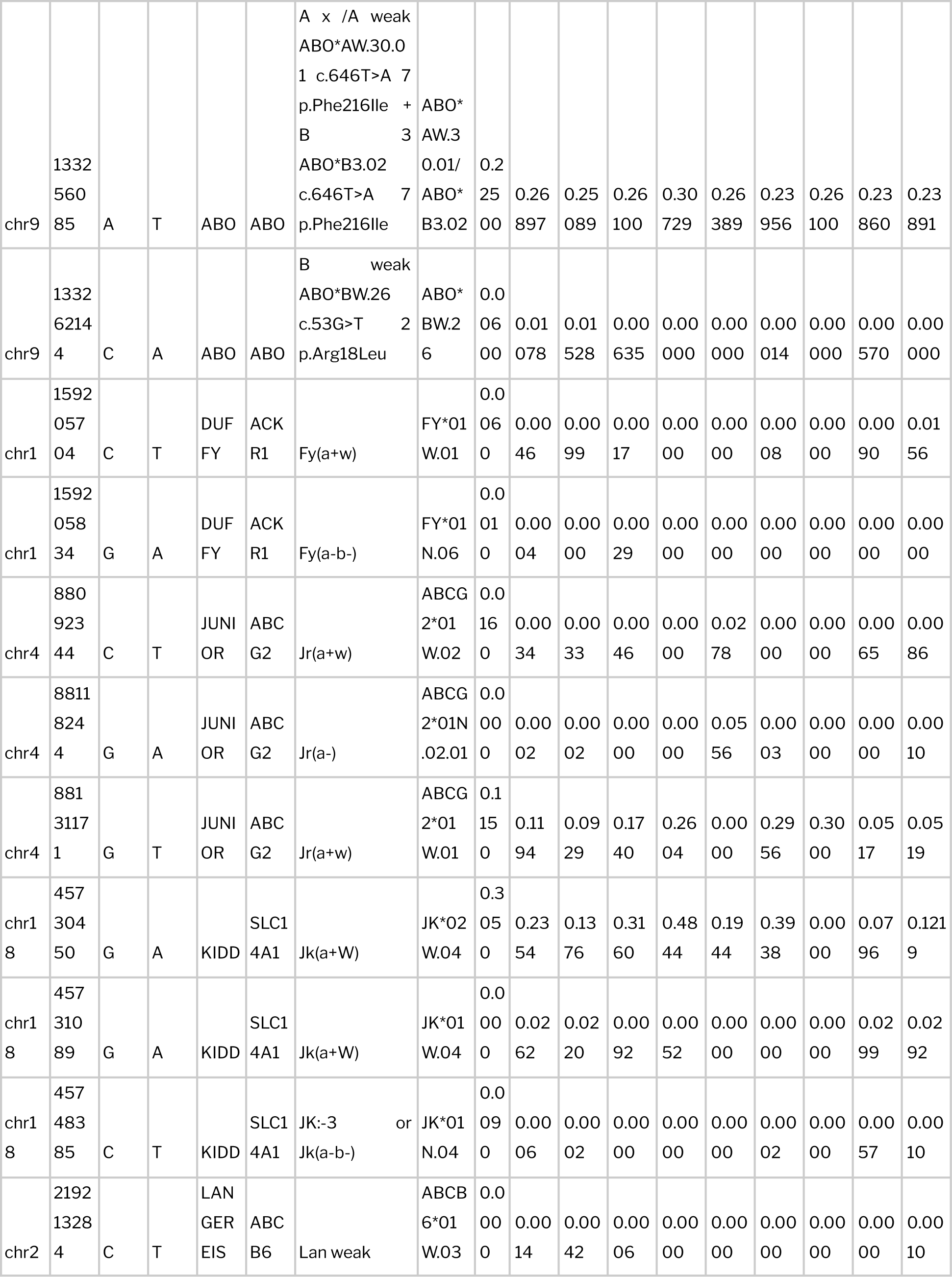
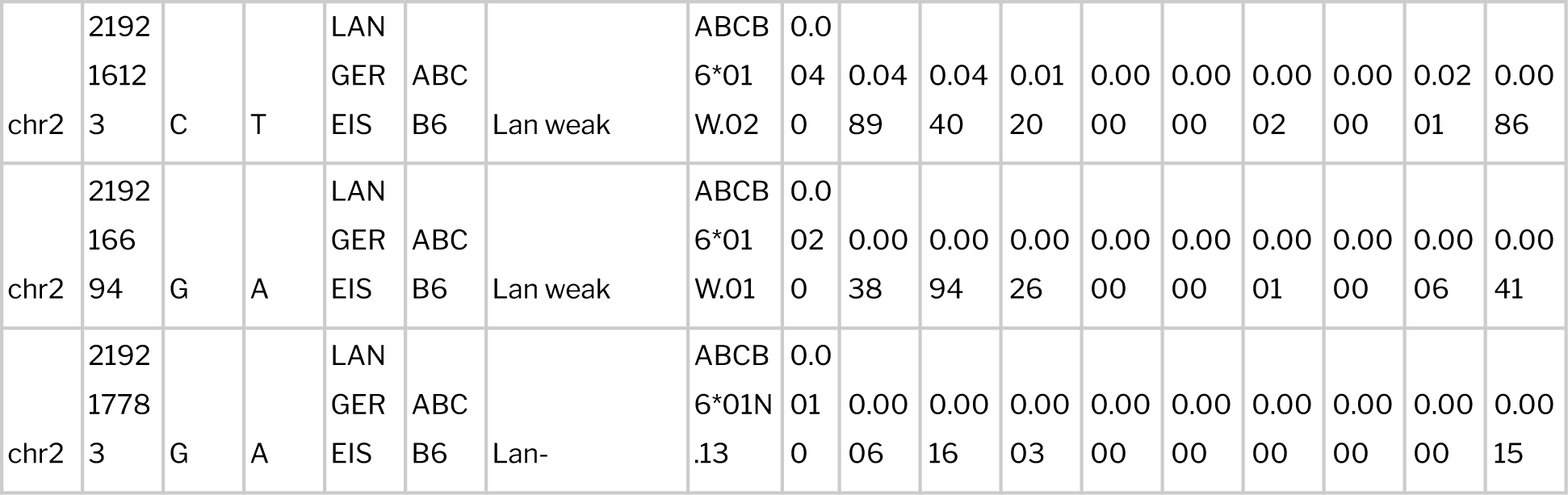
Compilation of weak, partial and null antigen frequency distribution among studies populations.

Anti-Jra antibodies of the Junior blood group system are known to cause transfusion reactions or hemolytic disease of the fetus or newborn. Interestingly, we identified a weak Jr antigen (ABCG2*01W.01 - c.421C>A; p.Gln141Lys), which is previously reported to diminish the expression of Jra antigen on the RBC membrane thereby leading to misinterpretation during serological testing {ref}.

The Kidd blood group poses several challenging transfusion scenarios. In addition to the typical acute hemolytic transfusion reactions associated with clinically relevant blood group antigens, weak Jk antigens are particularly notorious for triggering delayed hemolytic transfusion reactions (Lawicki et al., 2017; S.-S. Wang et al., 2021). Our study identifies 2 weak Jk antigens (JK*01W.04, JK*02W.04) and a null antigen (JK*01N.04).

In Langereis blood group system, 3 weak (ABCB6*01W.03, ABCB6*01W.02, ABCB6*01W.01) and 1 null antigen (ABCB6*01N.13) variant was identified. Lan is one of the high prevalence antigens found in almost all global populations. Anti-Lan antibodies are well known to cause delayed hemolytic transfusion reactions in adults as well as hemolytic disease in fetuses and newborns with varied levels of clinical significance (mild-severe).

### Rare blood phenotypes that lacks high prevalence antigens

High prevalence or high frequency blood antigens are those that occur in more than 99% of the population. Owing to their rarity they become difficult to identify using routine serological screenings. Patients lacking a high prevalence antigen possess a greater risk of alloimmunization while transfused. In our study dataset, 2 distinct rare blood group phenotypes lacking a high incidence of antigens were identified.

In the Diego blood group system, there exist three major high prevalence antigens - Dib, Wrb, and DISK. The rare Di(a+b-) phenotype was observed in 0.1% in the Indian populations. The Yt blood group system consists of 2 antithetical antigens - high-prevalence Yta and the significantly lower prevalence Ytb antigen (Van Buren et al., 2022). Upon analysis we found that the rare Yt(a-b+) phenotype was found in 0.29% of the population. Individuals possessing these phenotypes require antigen negative blood to avoid hemolytic transfusion reactions (HTRs). KANNO1 is a high prevalence antigen belonging to the recently categorized KANNO blood group system (Ohto et al., 2022). Lack of this high prevalence antigen has been previously reported among the East and South Asians. This rare KANNO-phenotype was found in 0.1% of the study dataset. To this day, there have been no documented instances of hemolytic transfusion reaction or hemolytic disease of the fetus and newborn resulting from anti-KANNO1 antibodies.

### Comparison of blood group profiles between India and global populations

Allele frequencies of all the blood variants filtered from the study were compiled and systematically compared with global population datasets. Initially, the distribution of frequencies of the filtered variants within the subpopulations of the 1000 Genomes and gnomAD datasets was examined. As shown in **Figure 3 and 4**, although the frequencies of a handful of the variants followed a similar range between Indian and Asian datasets, differences in overall distribution was also observed. Allele frequencies observed across other population datasets are tabulated in **Table 4**. Complete list of frequencies observed in the subpopulations is tabulated in **Supplementary Tables 3a and 3b.**

**Figure 3.**
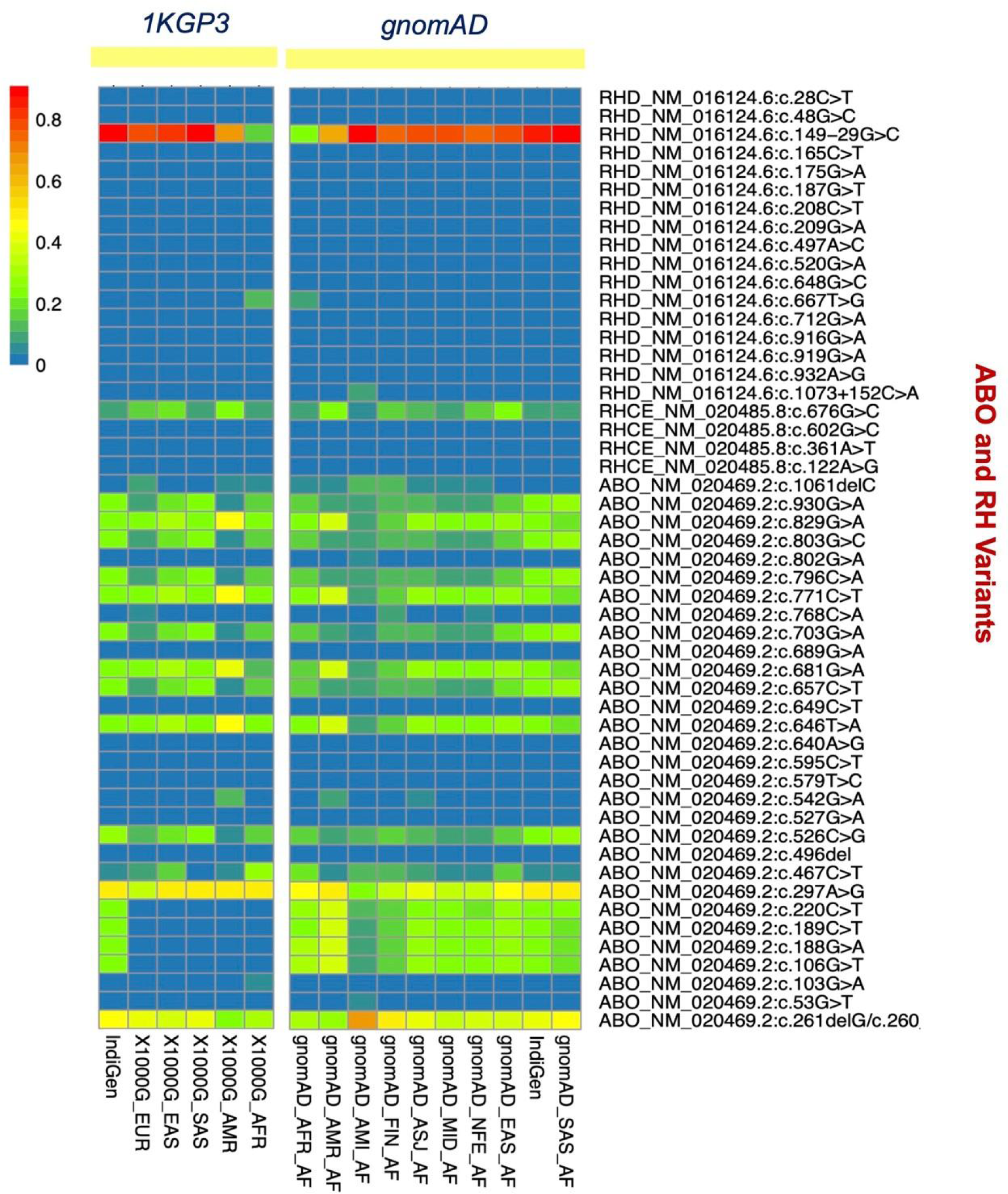
Heatmaps depicting the distribution of frequencies of the blood variants filtered in the study. **(A)** Frequency distribution of the 50 variants belonging to ABO and RH blood groups among various subpopulations of the 1000 Genomes dataset. **(B)** Frequency distribution of the 50 variants belonging to ABO and RH blood groups among various subpopulations of the gnomAD dataset.

**Figure 4.**
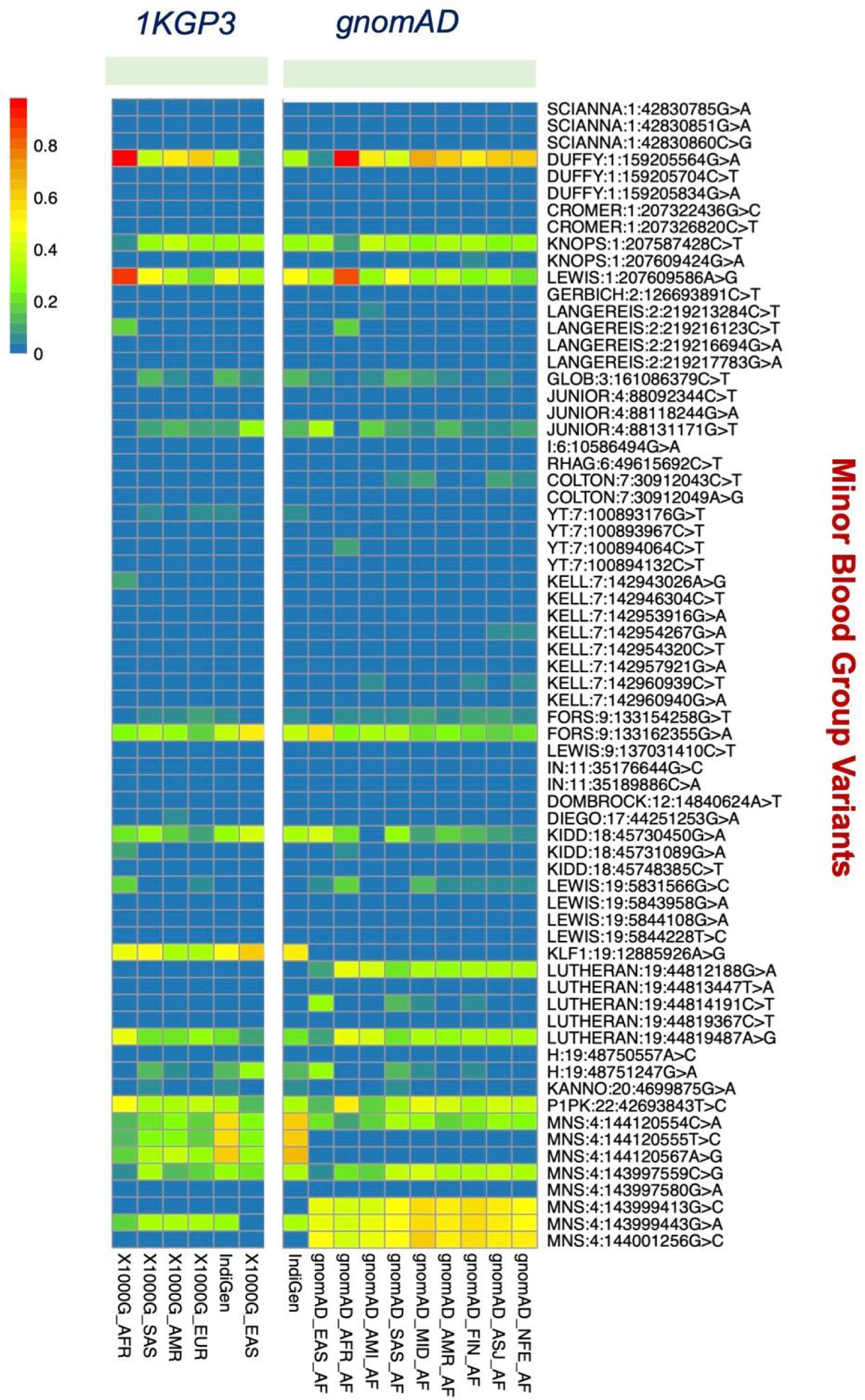
Heatmaps depicting the distribution of frequencies of the blood variants filtered in the study. **(A)** Frequency distribution of the 68 variants belonging to minor blood groups among various subpopulations of the 1000 Genomes dataset. **(B)** Frequency distribution of the 68 variants belonging to minor blood groups among various subpopulations of the gnomAD dataset.

**Table 4.**
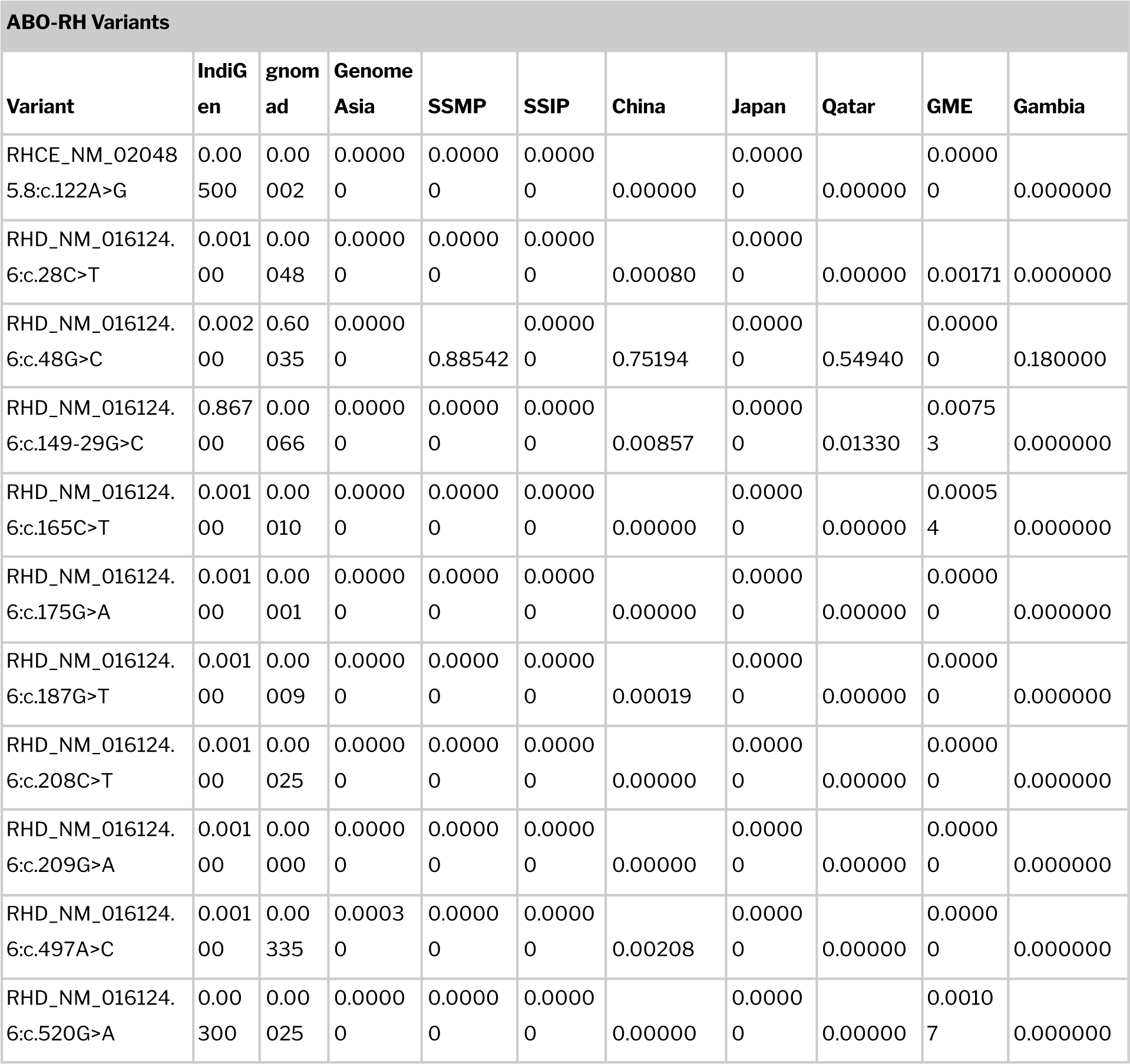
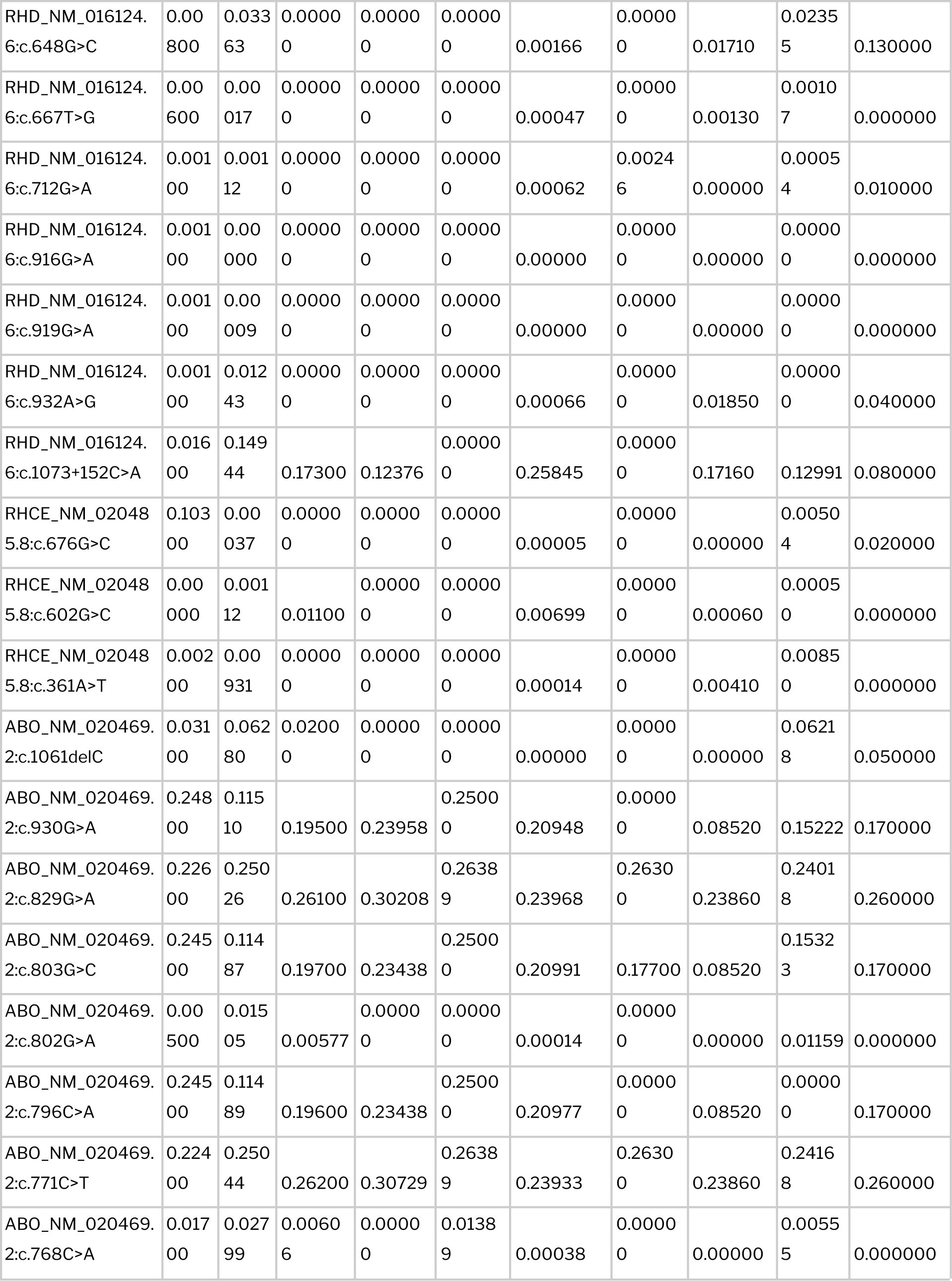
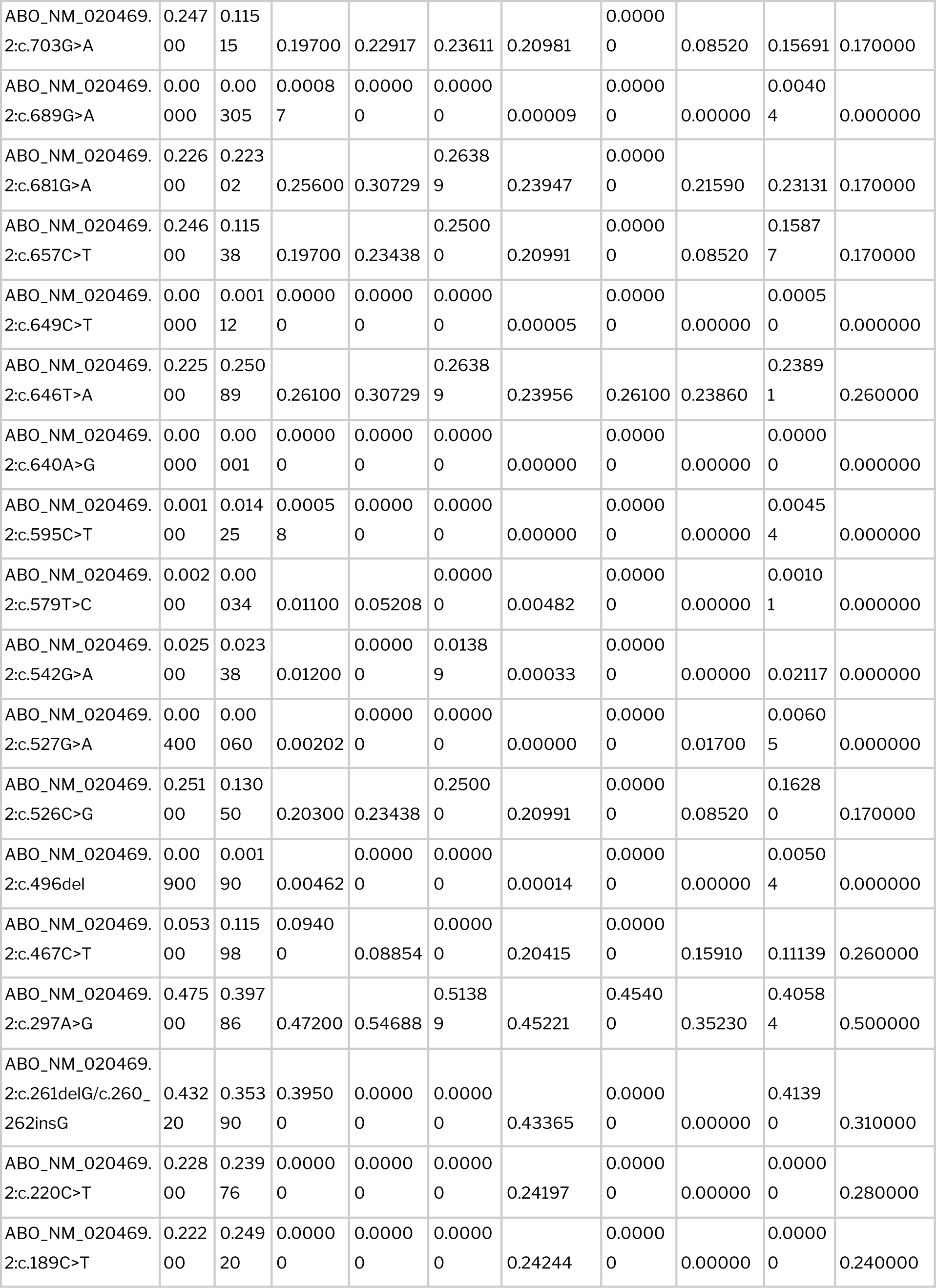
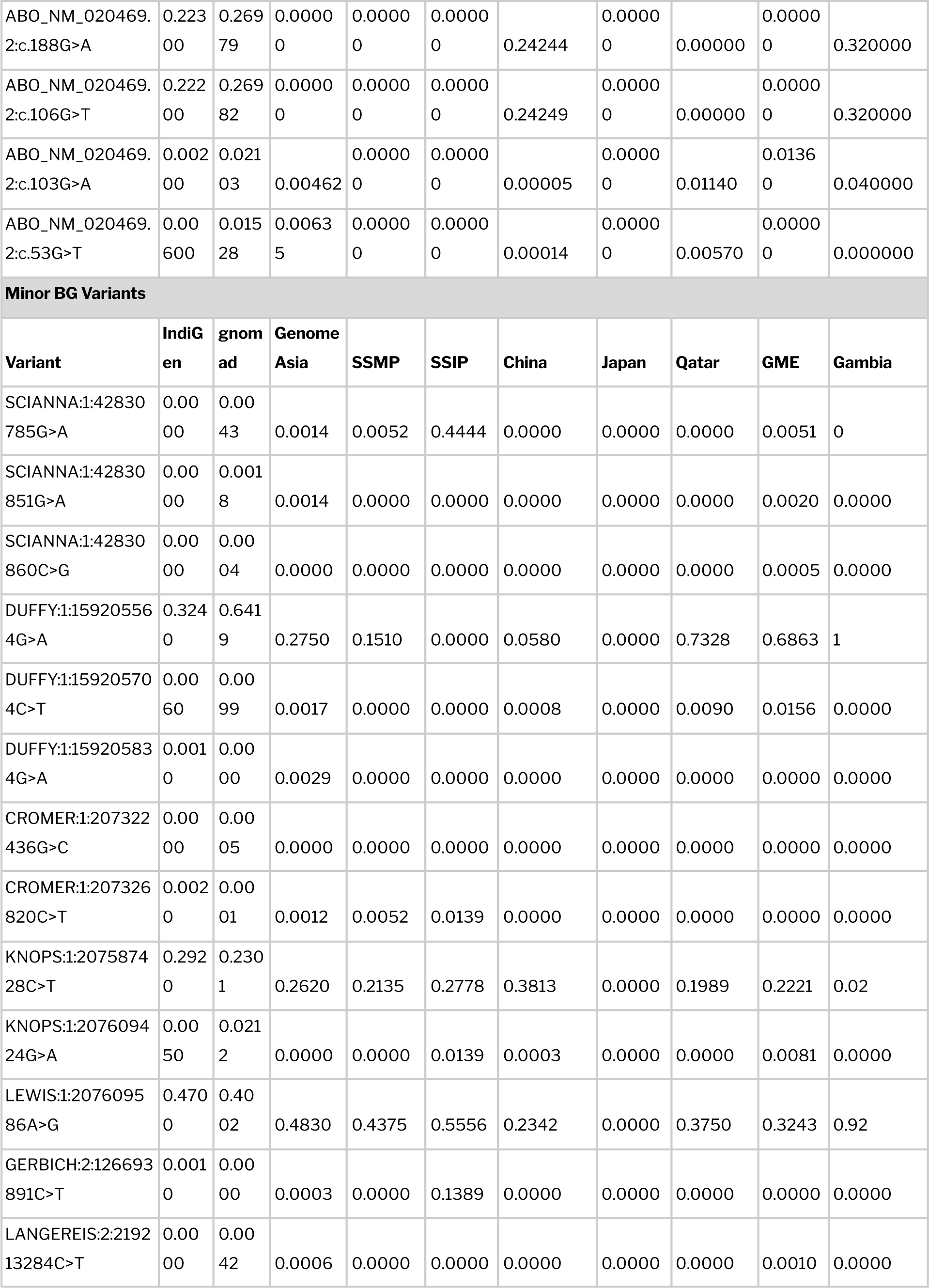
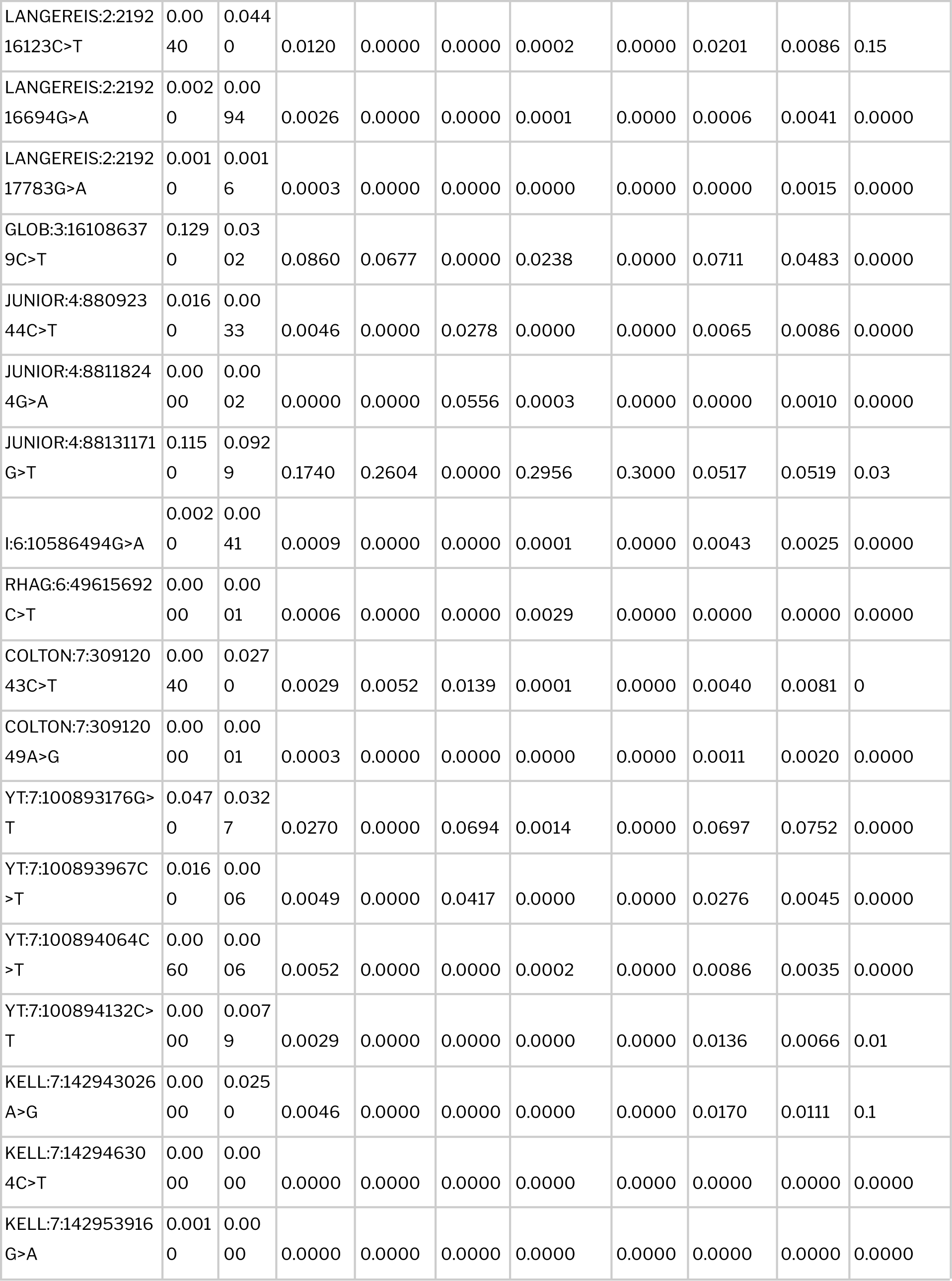
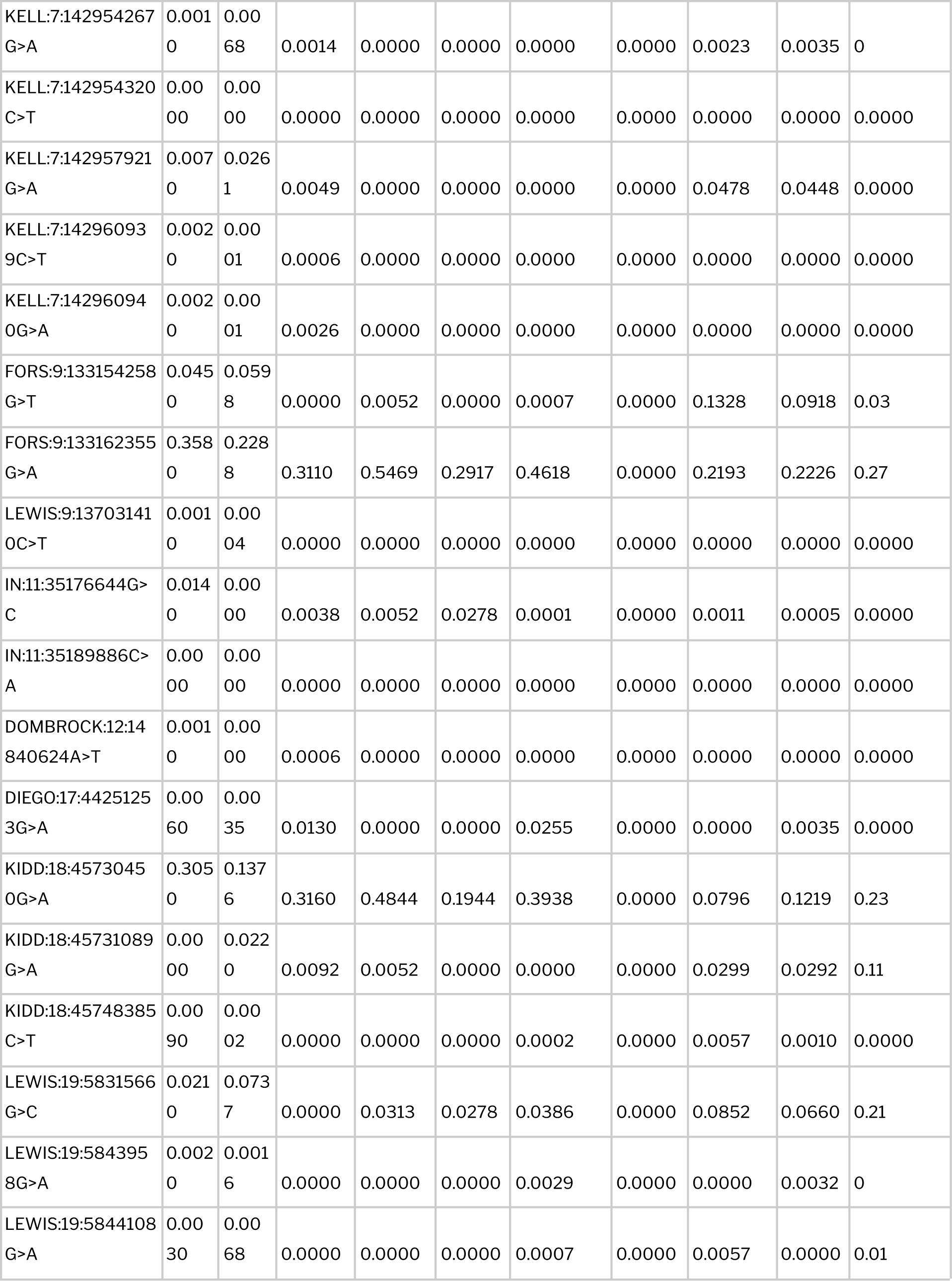
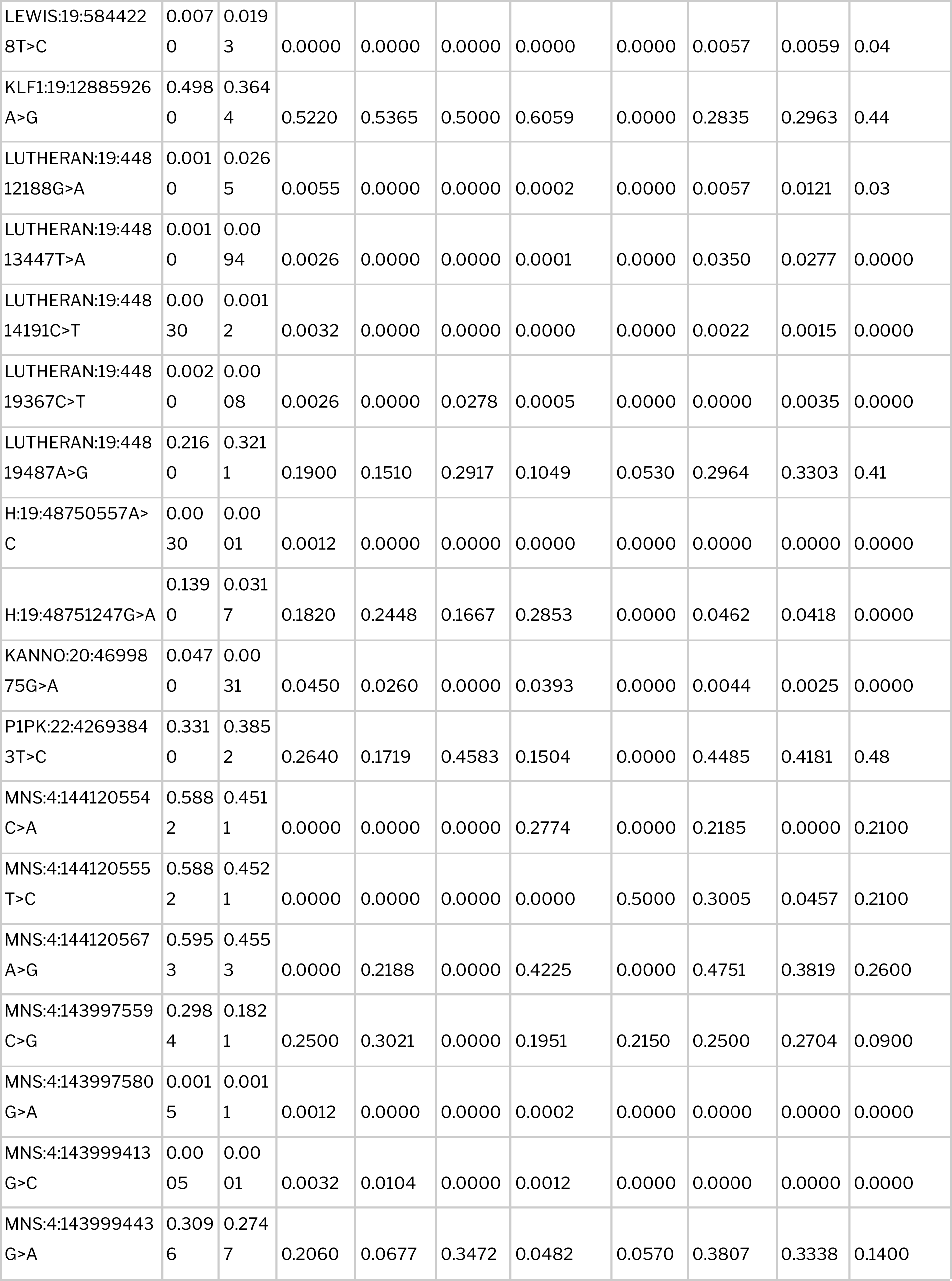
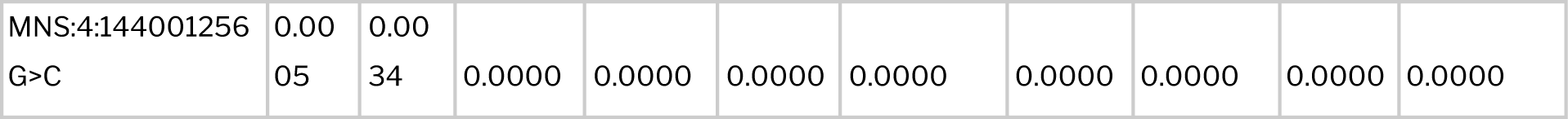
Overall distribution of blood variant frequencies across various global population datasets used in the study.

**Table 5.**
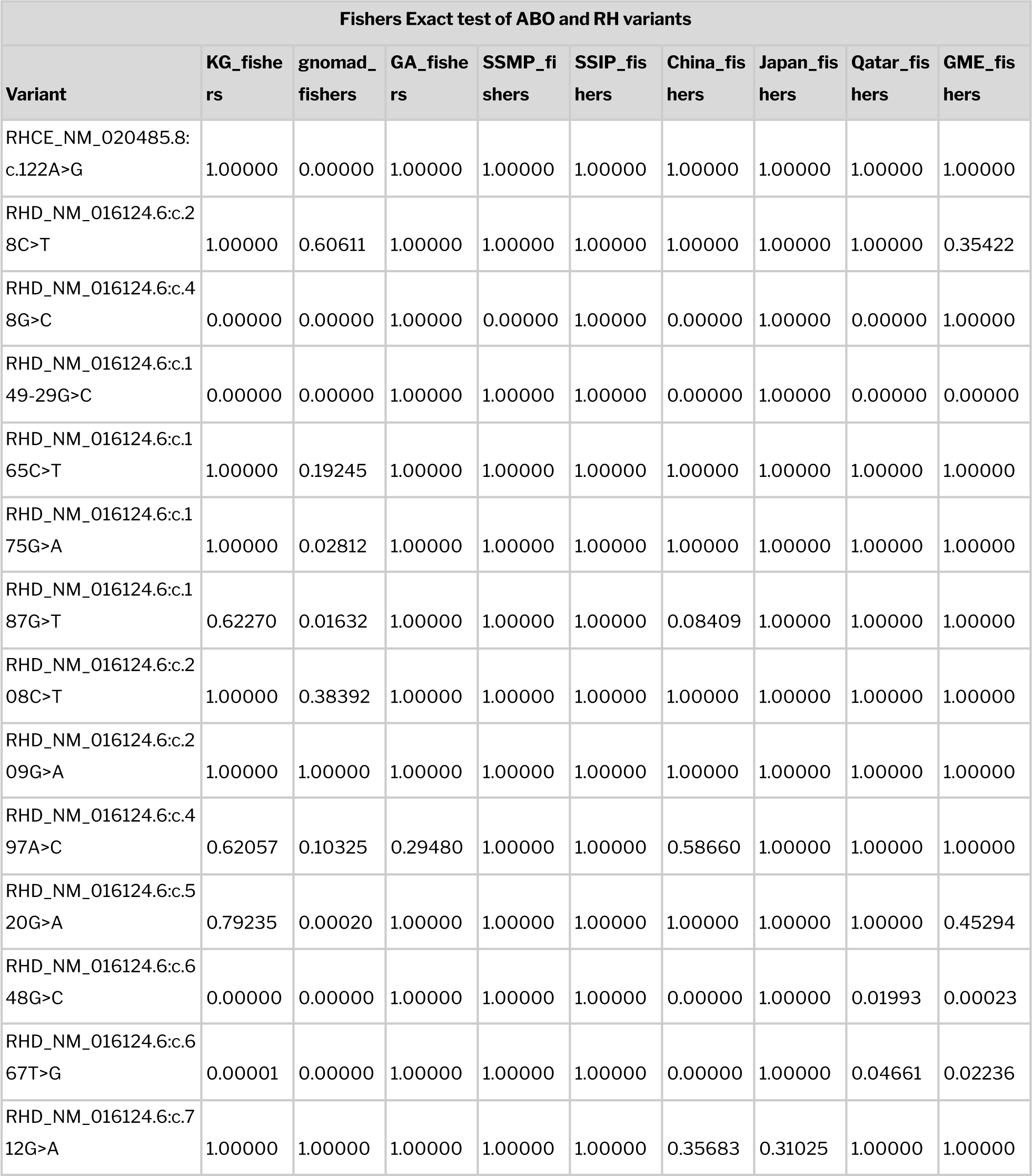
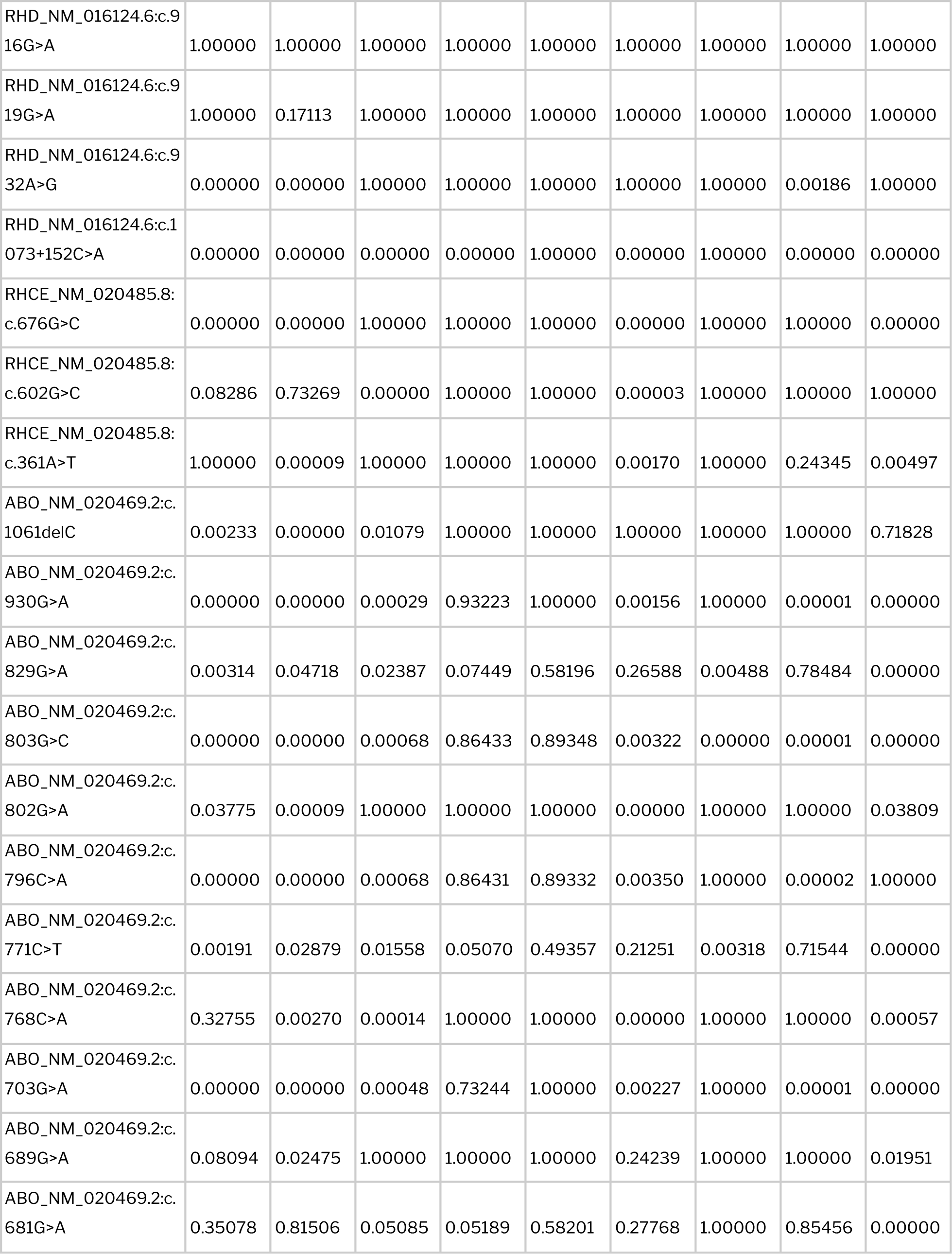
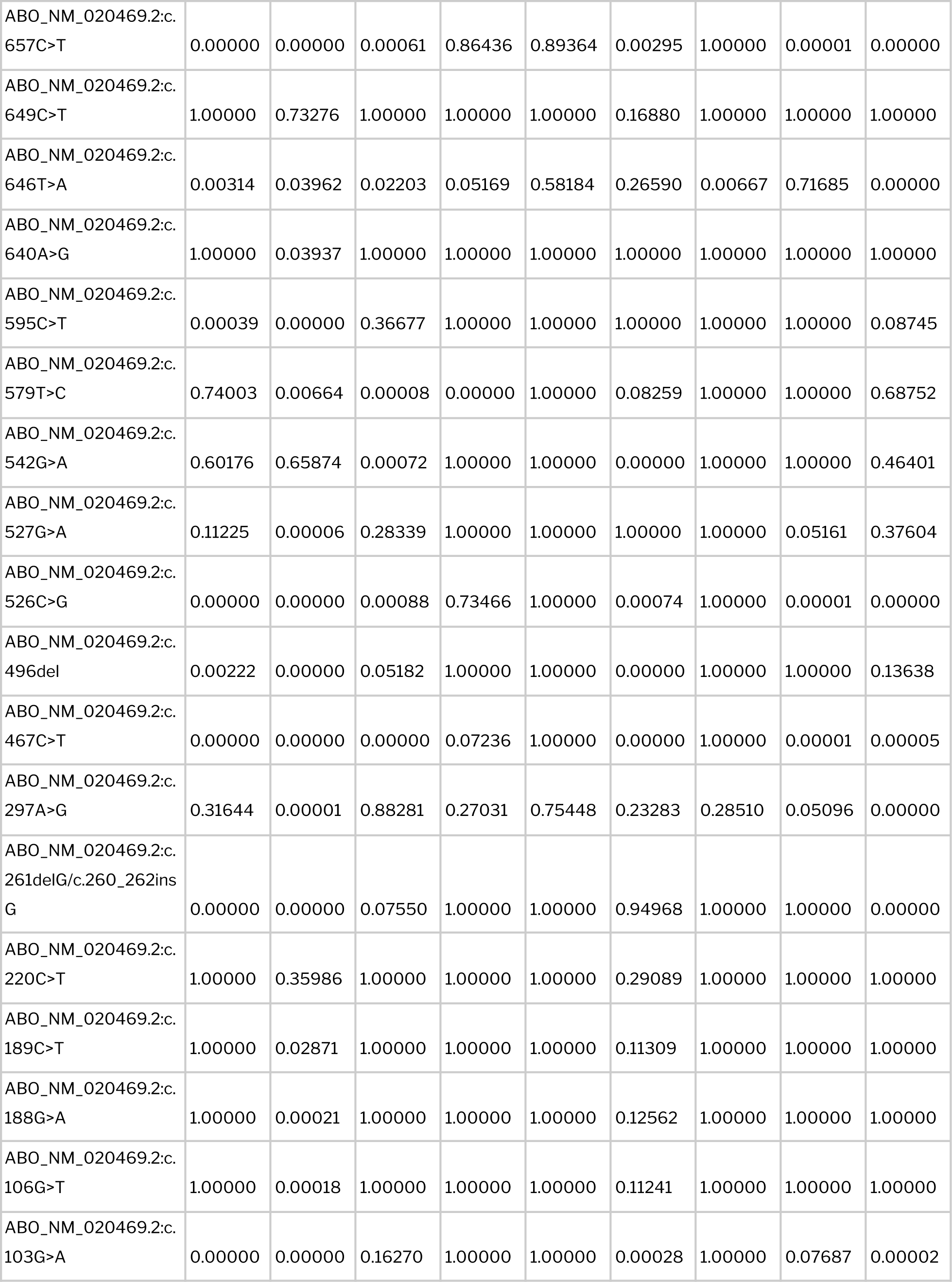
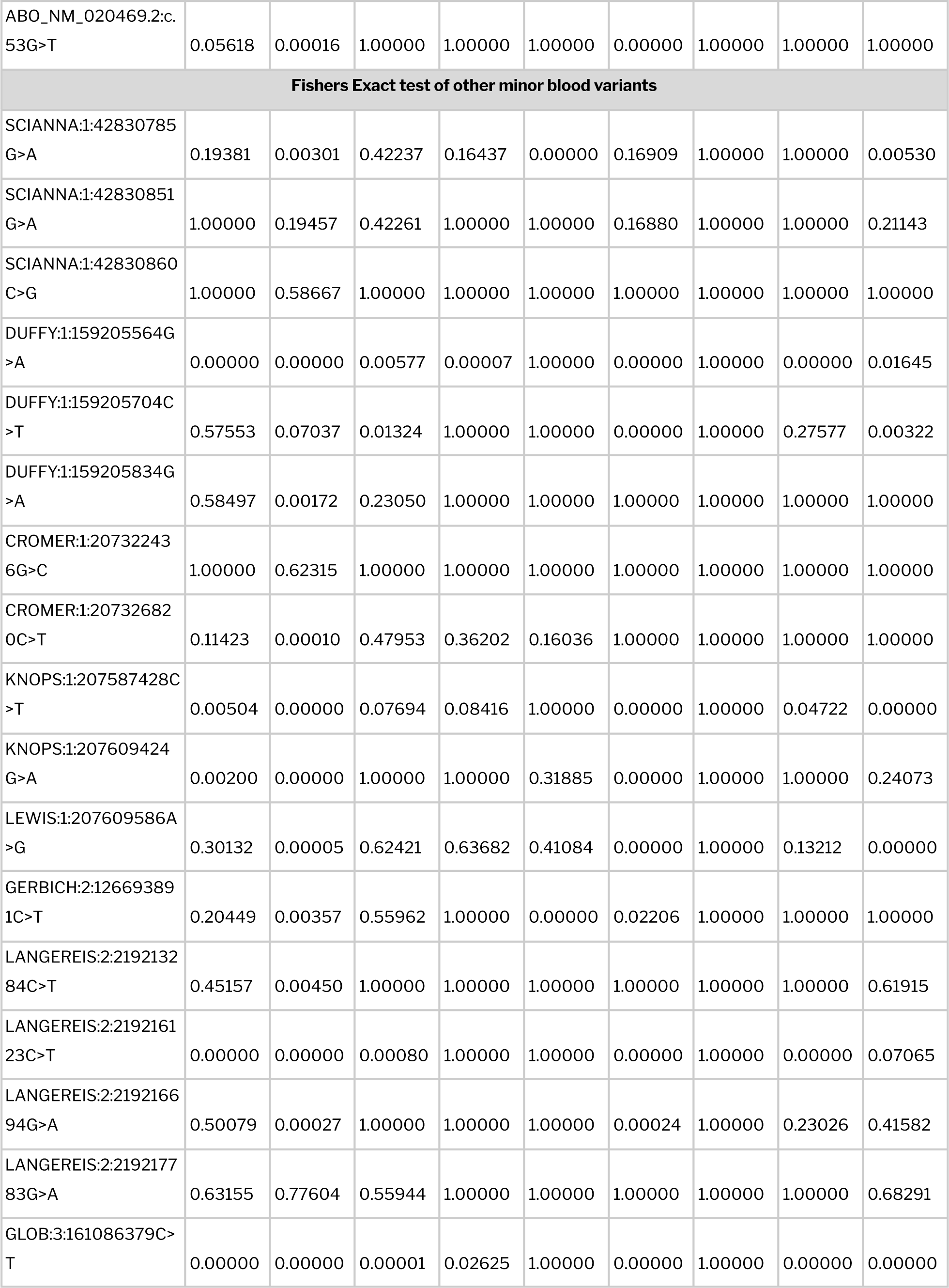
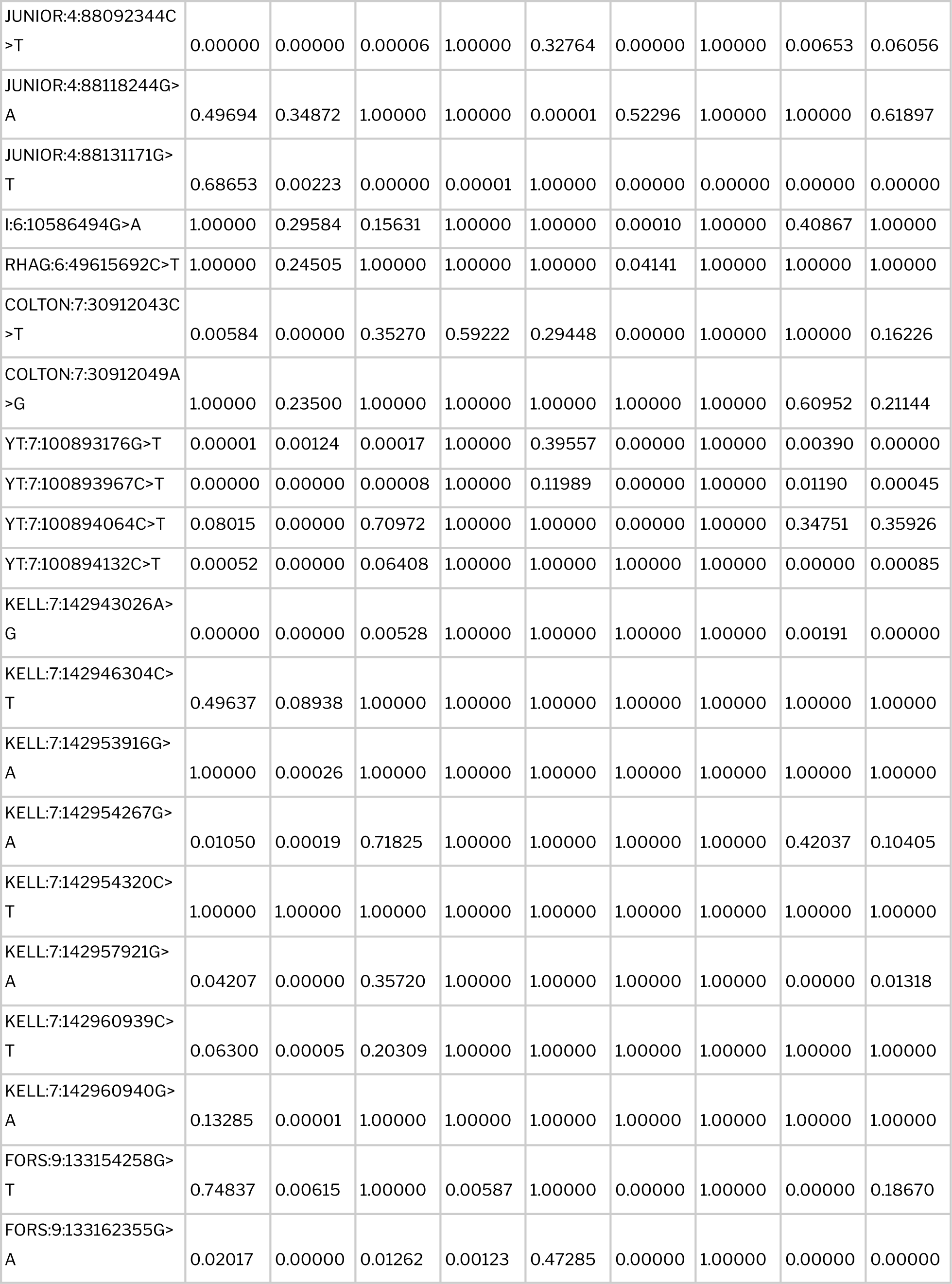
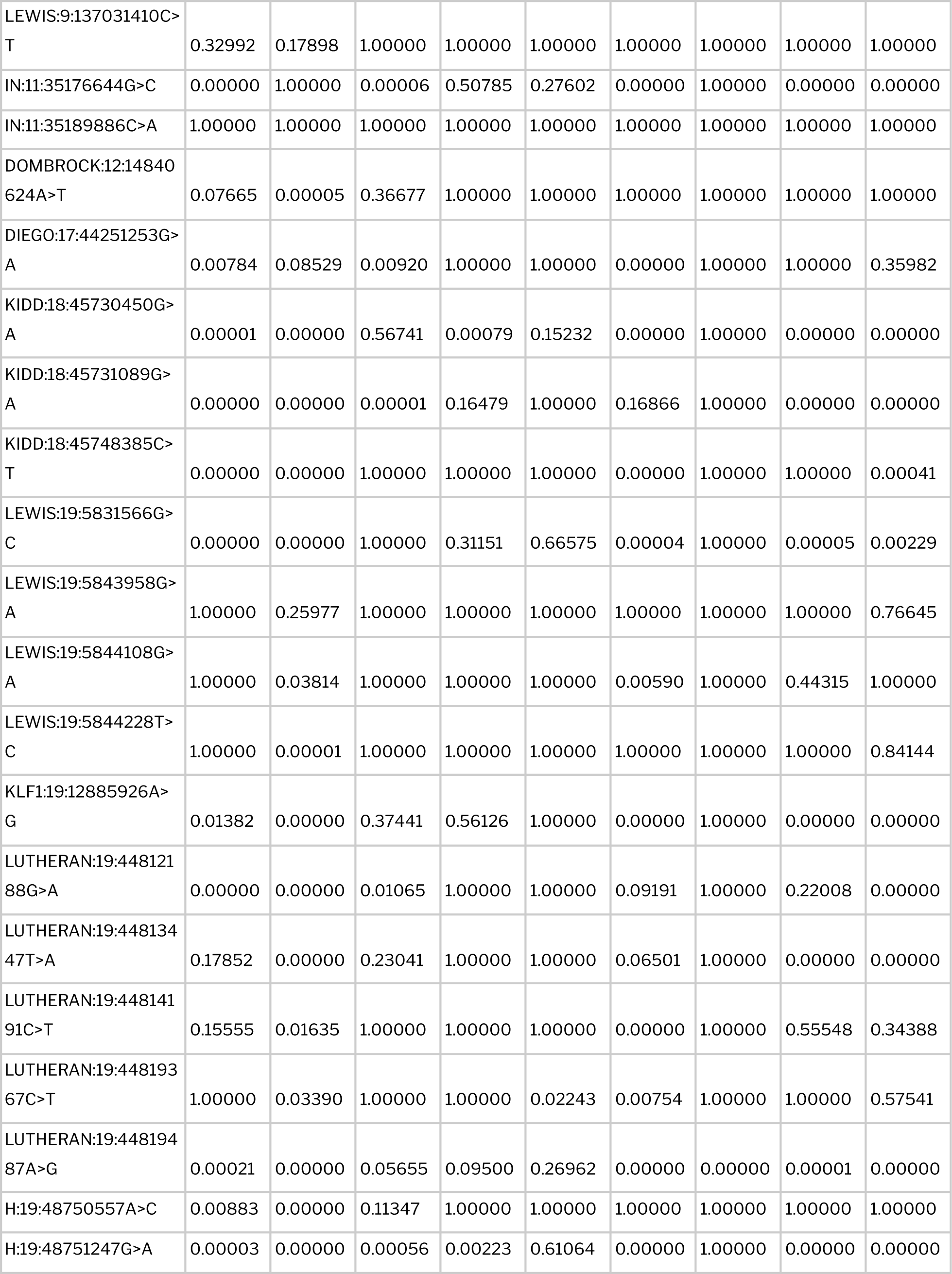
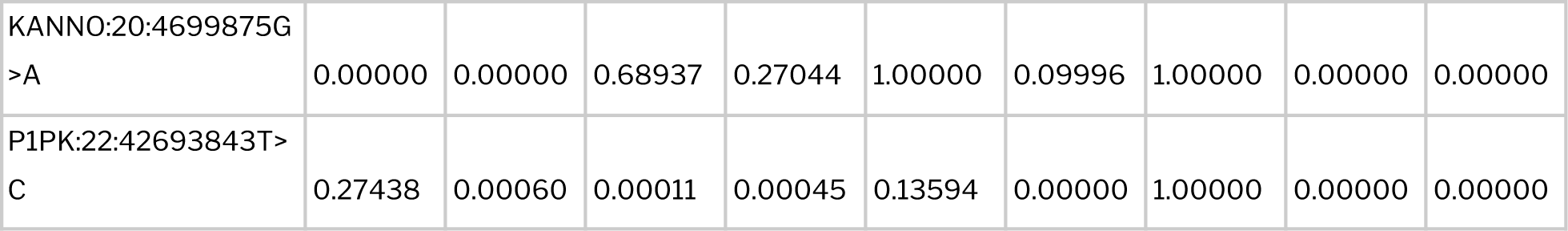
Tabulation of blood group variants which were found significantly distinct in the global populations along with their p-values.

Differences in the distribution of variant frequencies between India and a range of global populations namely Americans (gnomAD_AMR), Europeans (gnomAD_FIN, gnomAD_NFE), Africans (gnomAD_AFR, Gambian genomes), Middle Easterners (Qatar, GME variome, gnomAD_MID), Asians (GenomeAsia100K), East Asians (gnomAD_EAS, China, Japan), South East Asians (SSMP, SSIP) and South Asians (gnomAD_SAS) was analysed by performing Fisher’s exact test. Variants with p-value < 0.05 were considered significantly distinct.

One variant belonging to the Duffy blood group system *DUFFY:1:159205564G>A* responsible for encoding the Fyb antigen was found significantly different in almost all global populations. Duffy blood group gene, ACKR1 is highly polymorphic with allele frequencies greatly varying among different populations and ethnic groups (Höher et al., 2018; Howes et al., 2011; Shimizu et al., 2000). In compliance with previous reports, our analysis revealed a high prevalence of Fyb allele frequencies in European (60%), American (54%) and Middle Eastern (66%) populations. On the contrary, East Asians exhibited a very low frequency (7.7%) of Fyb allele as previously reported. Frequency of this allele in India (32%) was found comparable to other South Asian populations (gnomAD_SAS : 38%, 1KG_SAS : 35%)

A total of 26 variants belonging to 13 different blood group systems were found significantly distinct in India when compared to other South Asian populations. 2 weak RHD variants were found significantly higher in Indians. As mentioned earlier, these two variants RHD_NM_016124.6:c.520G>A and RHD_NM_016124.6:c.667T>G encoding weak and partial D antigens namely RHD Type 33 and RHD*DFV respectively are reported to form allo anti-D antibodies in carriers if transfused with D+ RBCs.

Further, on comparing the differences with other large populations based on their geographical locations including Africans, Americans, Europeans, Asians, South East Malays, East Asians, Middle East, Amish and Ashkenazi Jews we found that 37, 77, 77, 18, 26, 54, 23, 58 and 75 variants were found significantly distinct respectively. Complete list of variants that possess significantly different frequencies when compared with each global population are provided in **Table 4**. Schematic representation of the distribution of allele frequencies of significantly distinct variants is shown in **Figure 5**.

**Figure 5.**
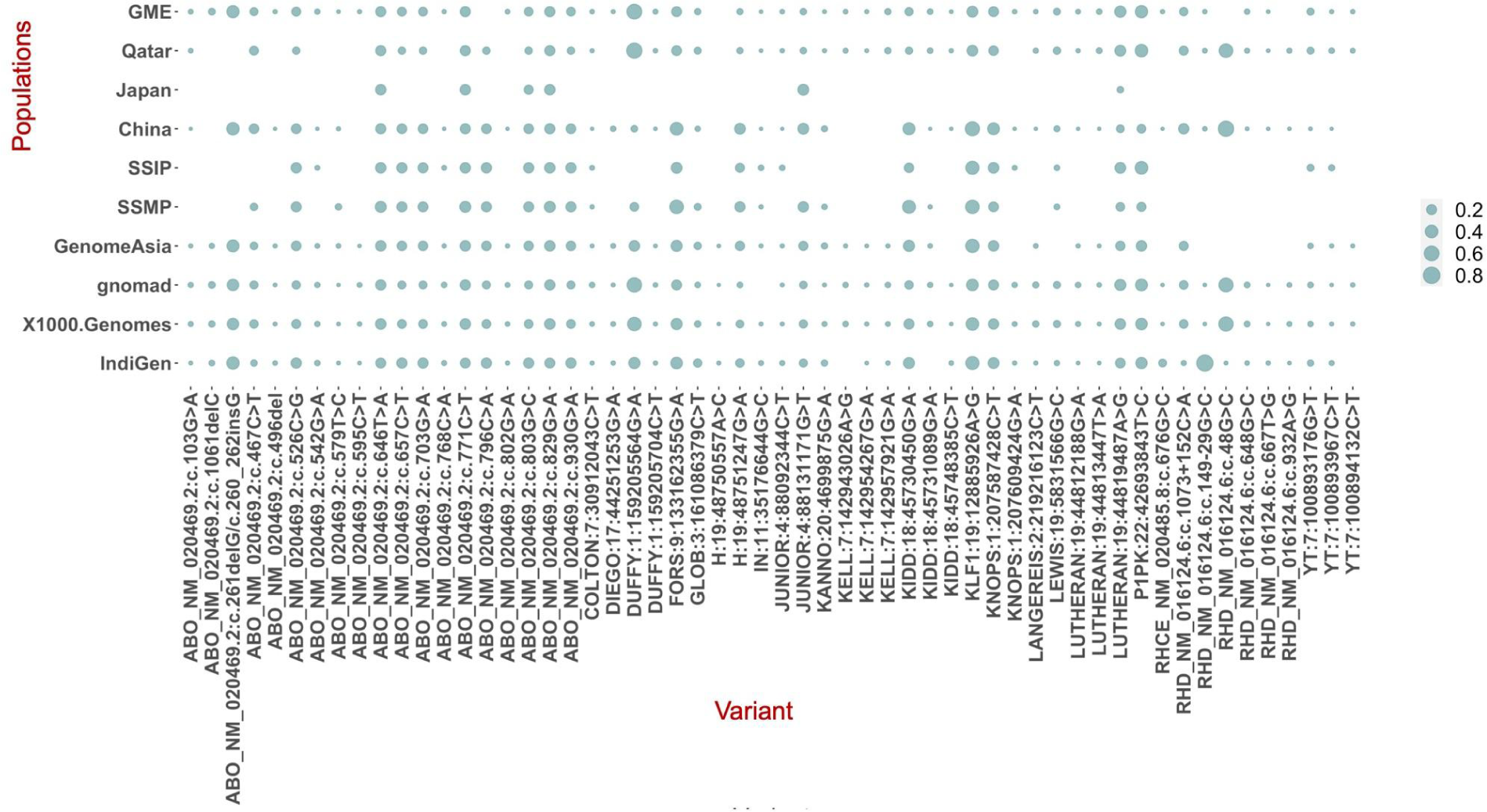
Bubble plot illustrating the schematic overview of the distribution of 56 variants across all blood group systems whose frequencies were found significantly distinct (p val < 0.05) in India in comparison to any of the other global populations.

## Discussion

Genomic analysis of 1029 self declared healthy Indian individuals revealed a total of 695 non-synonymous blood group associated variants mapping back to exonic and splice site regions. Preliminary level of analysis uncovered a subset of these variants which were found 2x enriched and depleted in the Indian population when compared with the 1000 genomes dataset. In addition, a range of weak, partial, null and potentially novel blood variants were also identified.

### Clinical relevance

In India, on analysing the prevalence and incidence of the diseases that systematically require transfusion, it was found that people with bone marrow failure (73%), leukemia (60.7%), end stage renal diseases (51.6) and blood disorders like hemolytic anemia (50.2) and hemophilia (25.6) were in higher need of blood units compared to others. In addition to specific disease conditions, trauma/surgery and obstetrics and gynecology are the two major fields which have huge demand for blood components. Recent estimates state that 6.6 and 3.6 million units of blood are needed to cater the surgical and obstetrics needs of the Indian population.

In December, 2012, The Hemovigilance Program of India (HvPI) was launched with the purpose of monitoring and preventing transfusion related adverse reactions (Bisht et al., 2020). A comprehensive 5-year study conducted by HvPI states that a total of 8162 cases of transfusion adversities were reported during the period. It is also found that 44% of all the complications caused by antigen mismatches were attributed to immunological hemolysis caused by other non-ABO alloantibodies. Such findings strongly emphasize the need to devise accurate ways of performing extensive blood group profiling which would in turn help in improving population specific donor registries in blood bank networks.

## Conclusion

Enhanced accuracy in blood group typing greatly reduces the risk of transfusion associated complications thereby leading to increased transfusion success rates. Our study serves as the first of its kind to provide a comprehensive compilation of the genetic landscape of blood group alleles and antigens in the Indian population.

## Data Availability

All data produced in the present work are contained in the manuscript

## Data availability

The Indian genetic variation dataset used in the study can be accessed at https://clingen.igib.res.in/indigen/

## Author Contributions

VS conceived and designed the project. MR and VS contributed in writing the manuscript. All authors approved the final manuscript. Authors acknowledge funding from CSIR India and Ministry of Higher Education, Malaysia. The funders had no role in the preparation of the manuscript or decision to publish.

## Supplementary Datasets

**Supplementary Table 1.** Locus genomic reference (LRG) coordinates of list of genes encoding human blood group, neutrophil and platelet antigens.

**Supplementary Table 2.** Tabulation of variant counts in each blood gene used in the analysis.

