## Supplementary figures and images for "Landscape of blood group antigens and alleles in the Indian population from whole genome sequences"

### Supplementary Figure 1

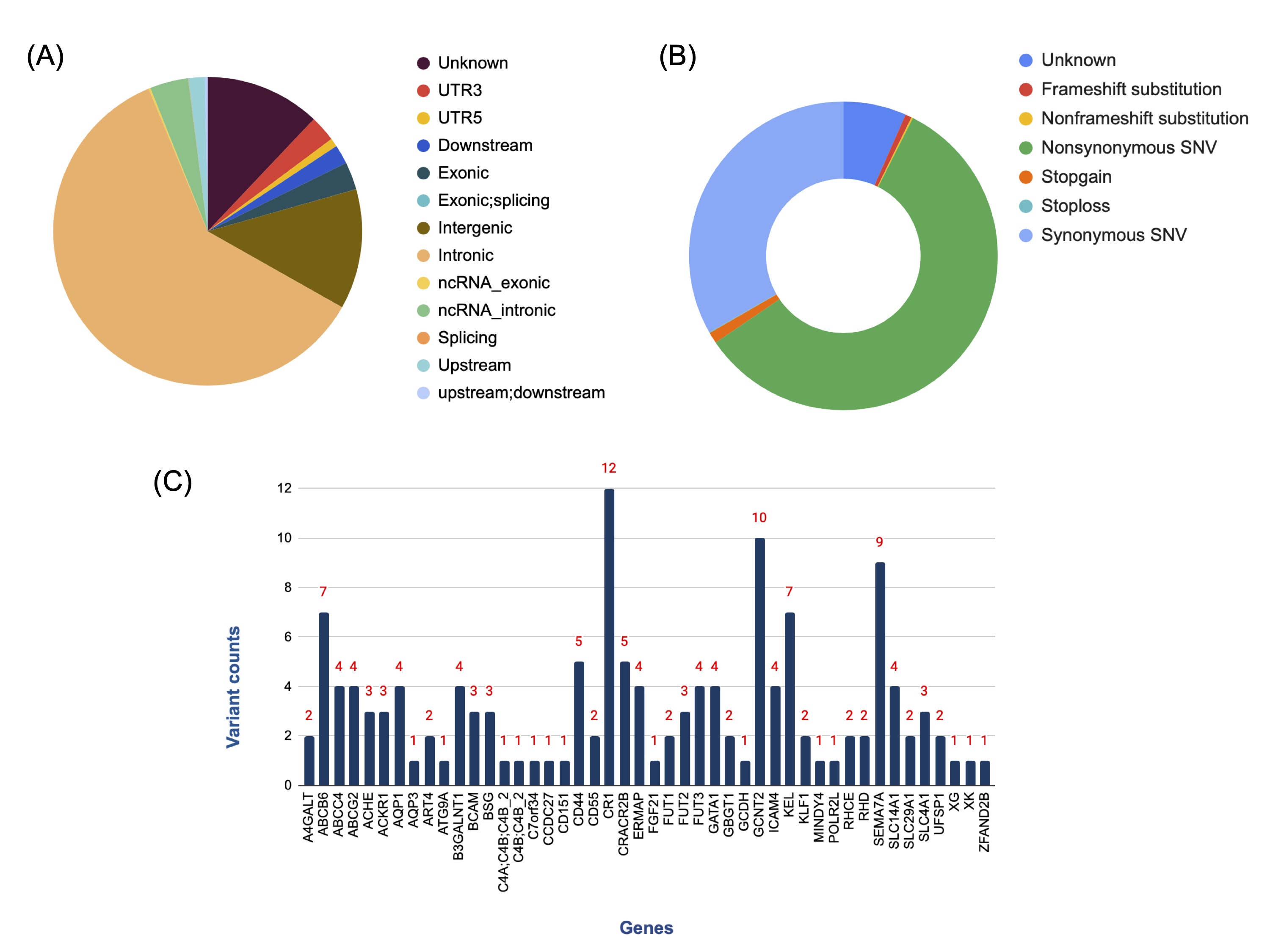
